# Framing of ultra-processed foods and associations with interests of actors quoted in UK news media, 2022-2023: a mixed-methods study

**DOI:** 10.1101/2025.08.15.25333779

**Authors:** Jennie C Parnham, Kiara Chang, Michael Essman, Deb Smith, Mike P Etkind, Maisie McKenzie, Milica Vasiljevic, Mark Petticrew, Martin White, Anthony A Laverty, Emma Boyland, Steven Cummins, Christopher Millett, Jean Adams, Eszter P Vamos

## Abstract

**Background:** There has been increasing attention on the health effects of foods classified as ‘ultra-processed’ (UPFs), with ongoing debate about the implications for policy. News media are known to influence public perceptions and perceived importance of action. Yet, there is a lack of understanding of how UPFs are framed in the media and how the UPF industry may contribute to this framing.

This study aimed to analyse the framing of UPFs in UK news media and trade press and explore the interests of individuals and organisations quoted in articles.

**Methods and Findings:** We systematically searched UK news media and trade press for articles which discussed UPFs in 2022-23. After screening, 188 articles were identified and analysed using thematic content analysis. Article text was examined for its position towards the concept of classifying food by processing-level (accepted/contested/mixed). Framing Theory guided the identification of UPF frames, which described patterns in how the problem was described, the proposed causes of the problem and suggested solutions. The interests of named actors quoted in the articles were classified using the International Committee of Medical Journal Editor’s (ICMJE) declaration of interests form and categorised into three groups: ‘Interests: UPF industry related’, ‘Interests: not UPF industry related’ and ‘No interests apparent’.

Half of the articles accepted the concept of classifying food by processing-level (52%), 19% contested it and 29% were mixed. Five UPF frames were identified. The frames ranged from presenting UPF harms as a systemic issue requiring governmental policy action (frame 1) to questioning the UPF concept (frame 4) and additionally highlighting the benefits of UPFs and proposing nutrient-based policy approaches including reformulation (frame 5). There were 116 named actors in the articles, of which 33% had UPF industry interests. Of this group, 45% were academics and 11% were industry spokespeople. Actors with UPF industry interests were more commonly cited in frames that contested the UPF concept (e.g. Frame 5: 73%) than accepted it (e.g. Frame 1: 10%).

**Conclusions:** This analysis found that half of the articles framed UPFs as an important, structural challenge to public health, proposing policy action. However, a fifth contested the UPF concept. Since academics made up the largest proportion of actors with UPF industry related interests and were more likely to be cited in frames that contested the UPF concept, this research highlights the need for closer attention to the interests of actors discussing public health topics in the media.

## 1 Introduction

Ultra-processed foods (UPFs) have received increasing media and public attention in the UK and globally^1^. The term was first formalised in 2009 by the Nova classification system^2,3^, which classifies food by their processing level (see Box 1), other similar classifications have since been developed^4^. This approach to dietary classification is distinct from other dietary measures as it groups foods by the extent and purpose of their processing rather than by their nutrient content or biological food group (meat, vegetables, etc). By considering the purpose of processing, as well as extent, the concept intrinsically identifies the role of large transnational corporations in global dietary transitions towards UPFs, with profitability serving as a major driver of their production and marketing^5^. In this way, the concept is unique in placing foods within a wider food system context when assessing the healthfulness of dietary patterns.

### Box 1

#### Nova Food Classification system^3^

**Group 1 – Unprocessed or minimally processed foods (MPF)**: Foods which have undergone no or minimal processing from their original or whole state from nature. Processing is performed to make the food edible or safe (including boiling, frying, pasteurising) to eat.

**Group 2 - Culinary ingredients**: Includes foods which are used in food preparation. They may be substances extracted from nature (salt) or be substances derived from group 1 foods (e.g. butter, olive oil).

**Group 3 - Processed foods**: Foods which have undergone processing using traditional methods (e.g. fermenting, salting) that combines MPF and culinary ingredients (e.g. salt, vinegar, sugar and fat).

**Group 4 - Ultra-processed foods (UPF):** Foods which have undergone extensive industrial processing and contain little or no whole foods. They may contain ingredients extracted from whole foods and artificial, non-nutritive ingredients.

Over the past 15 years, evidence on the health effects of UPF consumption has been building. Meta-analyses of longitudinal observational studies indicate an association between diets high in UPF and a range of non-communicable diseases^6–18^. Additionally, a cross-over RCT^19^ showed that UPFs increased *ad libitum* energy intake and weight gain over a two-week period compared to minimally processed meals matched for macronutrients. However, there is disagreement over whether evidence of the health impacts of UPFs is sufficiently convincing to justify policy action^20^. In particular, the relative lack of mechanistic evidence has prevented many authorities from acting, preferring to maintain policies driven by just the nutrient content and biological food groups. In addition, some have suggested that the Nova classification system is not precise enough for use in policy^20,21^. However, others have evoked the precautionary principle, arguing that the epidemiological evidence is already sufficient to warrant policy change^20,22,23^.

The media plays a crucial role in determining the perceived importance of an issue for both policymakers and the public and can have a significant impact on agenda setting and policy action^24,25^. Harmful commodity industries, such as the tobacco and alcohol industries, are known to use the media to influence public discourse to enable a more favourable policy landscape for their business interests^26–30^. Well-evidenced tactics include undermining scientific evidence, diverting attention away from the harms of their products and proposing solutions that avoid, delay or weaken regulatory change^31^. Additionally, it is well documented that industry funds scientific organisations, charities, interest groups and other actors to help promote these narratives and represent industry positions in the media^30,31^. As the Nova classification draws attention to the large corporations which produce and market UPFs globally^32^, there is a need to examine whether the discourse on UPFs in the news media is being influenced by the UPF industry (directly and/or indirectly through other actors).

The UK is an important case study, as there has been “*intensive public discussion and scientific debate*”^34^ over the concept of UPFs. The years 2022-2023 included significant UPF-related events in the UK, such as publication of a best-selling book on UPFs in April 2023^35^, a BBC documentary on UPFs in June 2023^36^, and the Scientific Advisory Committee on Nutrition’s position statement on UPFs in July 2023^37^. As discussions on potential UPF policy actions are ongoing, it is useful to analyse how the media frames the UPF concept to understand how it may be influencing public and policymakers’ perceptions of any potential actions. To date, one study has examined the use of UPF terminology in the Australian media, showing the concept is incorrectly presented in a third of articles^1^. Yet an in-depth analysis of how the concept is framed in the media, how particular framings gain prominence and potentially shape discourse, and the interests of the individuals or organisations who contribute to these frames is lacking. This study therefore aimed to analyse the framing of UPFs in UK news media and trade press (2022-2023) and the interests of actors quoted in these articles.

## 2 Methods

### 2.1 Research Design

We conducted a mixed methods content analysis of UK news media and trade press articles that discussed UPFs in 2022-2023.

### 2.2 Data source

We used Factiva to systematically search a selection of UK online and print news and trade press publications between 1^st^ November 2022 – 31^st^ October 2023. Factiva is a database of over 33,000 sources of news media (in print and online) globally^38^. The timeframe was chosen as there was an intensive focus on UPFs in the UK media during this period following a number of key events^9,35,36,39–43^, see **Figure 1** for a timeline. A list of publications was selected to provide a broad spectrum of national online and print news media and trade press in the UK. The list was chosen using national media-use figures^44^, previous media analyses^27,45^ and with guidance from our public partners (DS, MPE, MM, PS). Publications were categorised by their type, for example newspaper, online news, magazine or trade press (See Supplementary Table 2). Search terms were ‘*ultra-process*’, ‘ultraprocess*’, ‘UPF’, ‘food* near1 process*’* and *‘Nova’.* The search was restricted to articles which mentioned any of the search terms at least twice. An additional manual search of tonline news publications^44^ not included on Factiva (BBC News, Sky News etc, Yahoo News and Reuters) was conducted. The search found 961 articles, of which 332 identical duplicates (as identified by Factiva) were removed, therefore 607 articles were downloaded for full-text screening. The article title, text, date of publication, author, publication, publishing group and number of words were extracted to Excel. Initially, one researcher (JCP) conducted the full text-screening using the inclusion criteria (See **Table 1**). A second researcher (KC) then independently reviewed 20% of the articles. There was high agreement on article selection (85% relative agreement, Cohen’s κ= 0.71) with minor disagreements resolved through discussion, leading to a revision of the criteria which were applied to the remaining articles by JCP. Screening led to 72 further duplicates being removed; another 68 being removed for not meeting the article type or topic criteria and 301 being removed as UPFs were not the central focus (See **Supplementary Figure 1**). The final sample included 188 articles.

**Figure 1.**
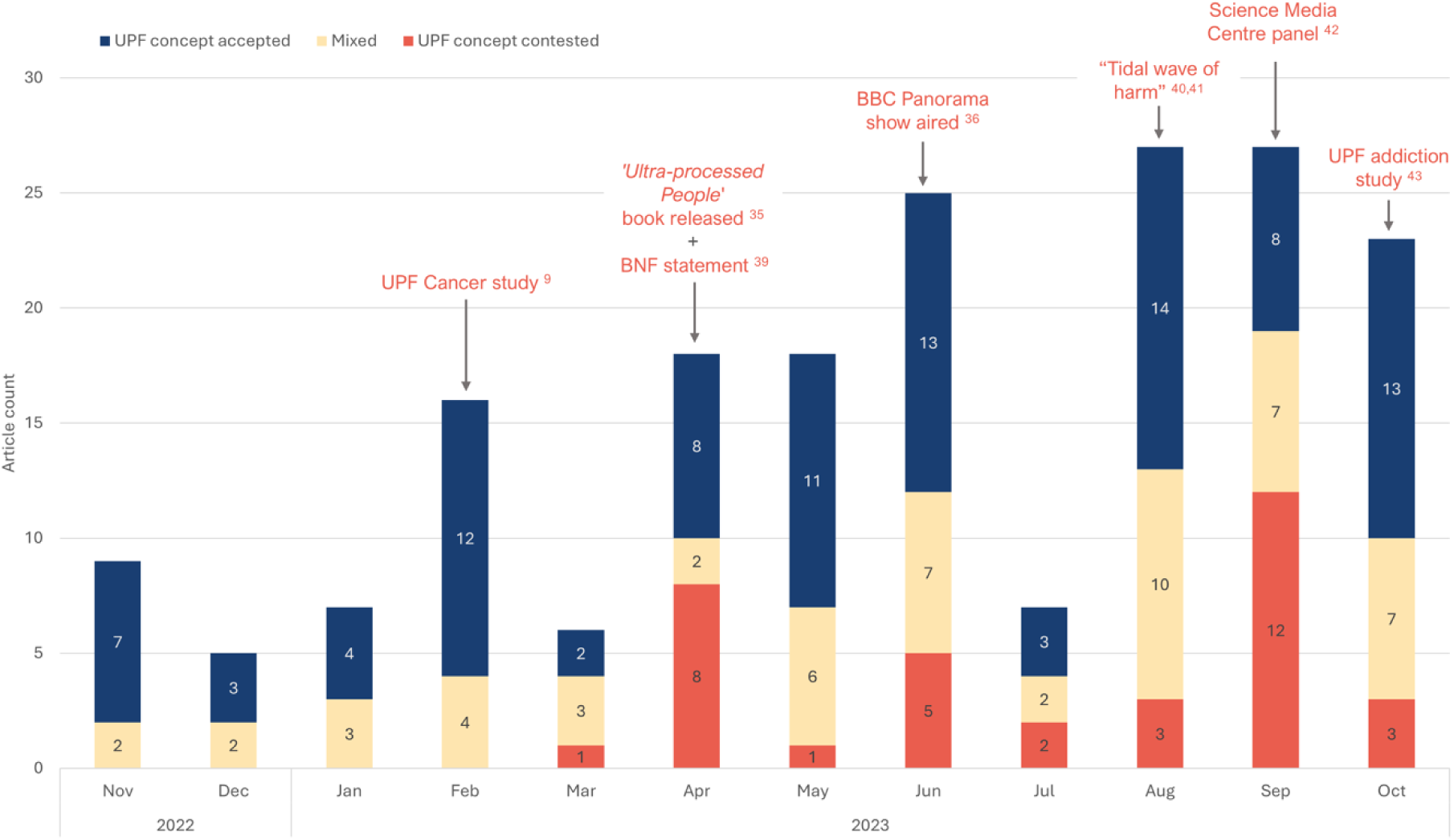
*– Number of articles discussing ultra-processed foods in UK news media and trade press between November 2022 to October 2023 per month by the overall article position towards ultra-processed foods.* Note: UPF – Ultra-processed foods; BNF – British Nutrition Foundation; Arrows show key events over the study period, see introduction and citations for more information.

**Table 1.** - Inclusion and exclusion criteria.

|  | **Include** | **Exclude** |
| --- | --- | --- |
| **Article type** | Original articles with editorial input over 100 words.<br>(e.g. news articles, feature articles, letters from readers, opinions) | News roundups<br>Radio or TV transcripts<br>Podcast, TV/radio programme, or event descriptions<br>Book best-seller lists |
| **Topic** | The concept, consumption or impact of ultra-processed foods (UPFs) including industries producing UPF. UK context or not country specific. | Articles where unhealthy foods were discussed generally but were not specific to UPFs<br>Articles where health behaviours, diet-related health or food policy were not explicitly linked to UPF.<br>Specific country-context outside of UK. |
| **Focus of article** | UPF were the central focus of article. | UPF were not central focus, they were mentioned but not expanded upon. E.g. UPF consumption was mentioned in a list of health-related behaviours |

### 2.3 Data Analysis

Analysis was conducted in two phases: qualitative and quantitative.

#### 2.3.1 Qualitative thematic content analysis

The thematic content analysis was conducted in NVivo 14^46^. Initially, a coding framework was developed iteratively using both an inductive and deductive approach^47^ after familiarisation with the data (JCP). The framework was structured around answering three questions.

First, codes were generated to assess the article’s discursive position towards the concept of classifying foods by the extent and purpose of processing, hereafter referred to as the ‘UPF concept’. Text that implicitly or explicitly accepted the concept was coded as “UPF concept accepted”, whereas “UPF concept contested” was coded to text which explicitly or implicitly questioned this concept (See **Supplementary Table 1** for a detailed description of these codes). The balance of these codes across each article was assessed and articles were given an overall categorisation as either “UPF concept accepted” or “UPF concept contested”, as appropriate. Articles were categorised as “Mixed” if both codes were present within the article.

Second, codes were generated to identify distinct ways that UPFs were framed across the data, informed by Framing Theory^27,48–50^. Entman proposed that “*to frame is to select some aspects of a perceived reality and make them more salient*”. This is done to promote a particular *problem definition* (hereafter: problem)*, causal interpretation* (hereafter: cause), *moral evaluation* and *treatment recommendation* (hereafter: solution) of an issue. Frames were differentiated by the themes that were included (or omitted) in a frame and where responsibility was placed (e.g. individual, government or industry). More than one frame could be present in an article, where articles presented opposing viewpoints on the topic. Public health and commercial determinants of health literature informed the naming of the frames^51–55^ (further details are presented in the Results and **Figure 2**).

**Figure 2.**
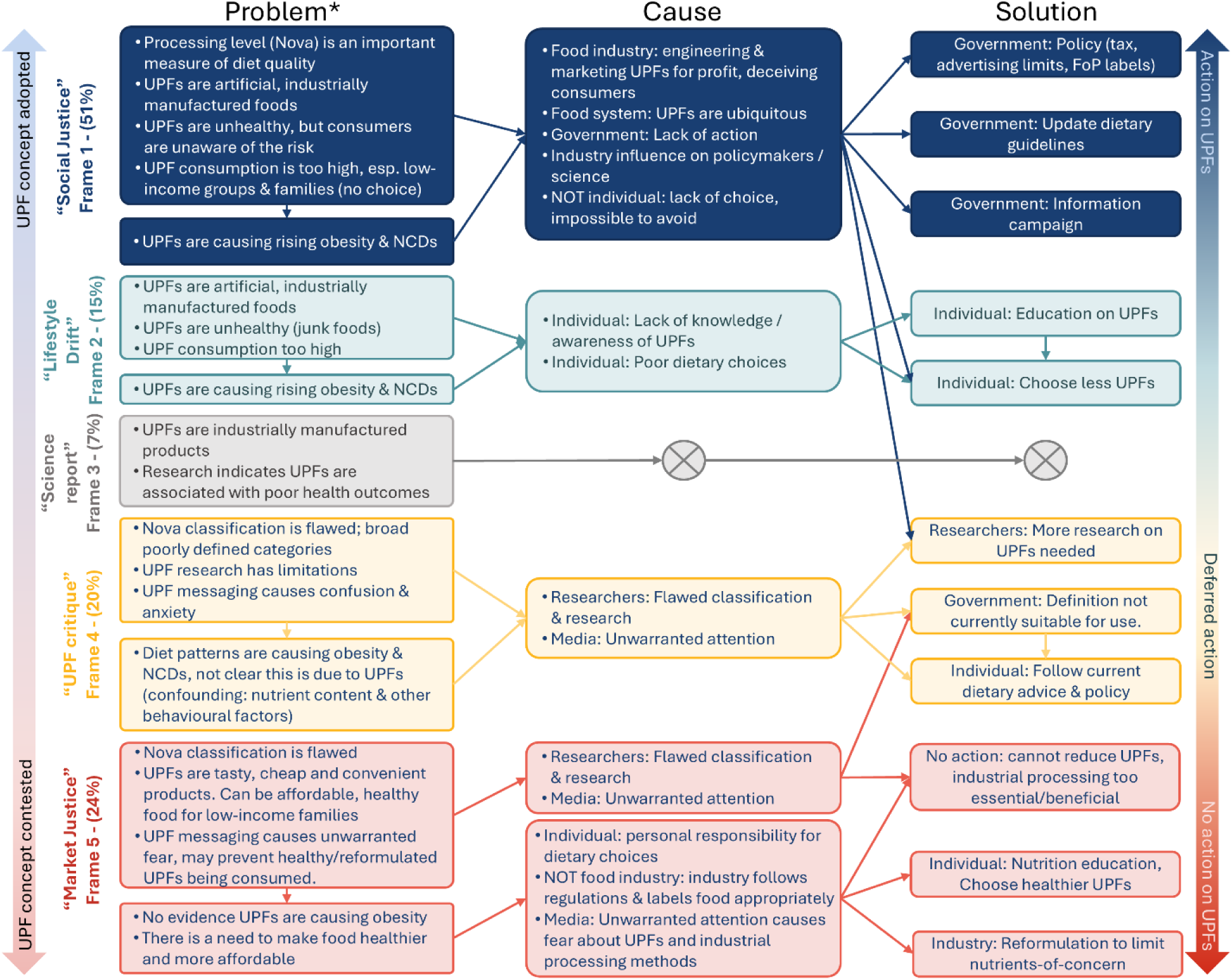
*– An overview of the five UPF frames derived from articles in UK newspapers.* *See **Supplementary Table 5** to **Supplementary Table 8** for a detailed overview of the frames. Note: Grey cross shows where causes and solutions were not proposed; More than one frame could be present in each article, therefore percentages add to over 100%; UPF = Ultra-processed foods; Esp. = especially; NCD = non-communicable disease; FoP labels = Front-of-package labels.

Third, codes were generated to identify named individuals or organisations in each article, referred to hereafter as ‘actors’, who were quoted in articles. Each actor was assigned a case in NVivo so their contribution across multiple articles could be assessed.

The initial coding framework was further refined after discussion with the research team. A second (KC) and third researcher (ME) independently read a random sample of articles (10%) and undertook independent coding. Key patterns in the framing of UPFs in the data were discussed among researchers (JCP, KC, ME) to check for consistency and reliability of the frame identification. Furthermore, our public partners (DS, MPE, MM, PS) individually read eight articles to further validate the frames. After discussion to resolve any discrepancies, the definition frames were refined. The final coding framework was applied to each article.

#### 2.3.2 Analysis of actor interests

In the data, 116 quoted actors were identified. These were extracted to identify their ‘interests’. For this analysis, the International Committee of Medical Journal Editor’s (ICMJE) disclosure form informed our definition of ‘interests’, which states that ‘*all relationships/activities/interests*’ with ‘*profit or not-for-profit third parties whose interests may be affected*’ within the past three years should be disclosed^56^. This includes research funding, royalties, stock interests, contracts, consulting fees, speaking/travel honoraria and participation or fiduciary roles in committees, boards or advocacy groups (paid or unpaid). Publicly available sources of information were used to identify interests, including institutional websites, academic papers, LinkedIn, scientific/advisory board meeting notes, organisation’s websites and annual reports. Google was used to search for the actor’s name (in quotations) with search terms (in quotations) including “profile” and a range of synonyms for “disclosure of interest” (see **Supplementary Table** 3). A consistent search formula that included quotations and a plus sign (“*actor name” + “search term”)* ensured the results were not personalised. Where an actor was indicated as acting in their capacity as a spokesperson for an organisation (industry, government, or non-government organisation (NGO)), their interests were determined at the organisational level. The ICJME’s definition states only interests within three years need to be declared, therefore 2019 was chosen as a cut-off for all interests, as it is three years before the start of the study period (2022-23).

Interests were further analysed (including through additional searches) to identify whether they were related to the UPF industry. For the purpose of this analysis, only market-leading UPF manufacturers were defined as ‘UPF industry’, using the same approach as previous studies^57^. First, if the company appeared on Slater et al’s^57^ list of 150 leading global UPF corporations, the interest was defined as ‘UPF industry related’. If an identified interest was a UPF manufacturer but was not in the top 150 global UPF companies, Euromonitor International’s Passport database^58^ was used to check if the company was in the top 10 companies by market share for their product in the UK or Europe, in alignment with Slater *et al.*’s methodology^57^. A list of companies defined as ‘UPF industry’ in this analysis are listed in **Supplementary Table 4**. Organisations reporting any funding from these UPF corporations (e.g. trade associations, charities or research consortiums) were also coded as ‘UPF industry related’. This is to capture the potential use of industry interest (or ‘front’) groups which have been previously reported to represent industry positions in the media^30,31^. Individuals or organisations who had declared interests that met the ICJME’s criteria but were not linked to the UPF industry (as defined for this analysis) were categorised as ‘Interests: not UPF industry related’. This included food/nutrition advocacy groups (with no links to UPF industry), those who received royalties from a food/nutrition book or received funding from non-UPF industry sources (e.g. medical companies). Those with no declared interests, or interests that did not meet the ICJME’s definition were categorised as ‘No interests apparent’. This label was used to reflect the fact that it is not possible for all interests to be identified through publicly available sources of information. The interest groups were then merged back into NVivo.

#### 2.3.3 Quantitative content analysis

The codes generated in the qualitative analysis were used for quantitative content analysis. First, the distribution of articles by their overall position towards the UPF concept (accepted, mixed or contested) was calculated and summarised by month. Second, the distribution of article position towards the UPF concept by publisher type (newspaper, trade press etc.) was also calculated. However, low expected frequencies in over 20% of cells precluded the use of χ² test. Lastly, the frequency that article text coded to an actor was also coded to a UPF frame was calculated (at the article-level) and summarised by the actors’ interest group, using a χ² test to determine independence of association. Quantitative analysis was performed in Python (v 3.12.7).

### 2.4 Public involvement

Five public partners were involved throughout the research project (DS, MPE, MM, PS, SJ). They contributed to the research proposal, article search strategy, theme development and interpretation and contextualisation of the results, and writing and revision of the paper.

### 2.5 Reflectivity statement

The research team consisted of a mix of academic and public partners. The academic team has broad experience in public health, epidemiology, food systems and policy evaluation, including UPFs and the commercial determinants of health. The public partners have previously contributed to various research projects in the fields of health and policy. Consequently, the team brought extensive expertise in public health to the project, informing our understanding of the themes identified in the paper. However, the group holds diverse perspectives regarding the classification of foods as UPFs, the strength of the evidence, and the need for policy change.

## 3 Results

### 3.1 Description of the sample

Across the study period, 188 articles met the inclusion criteria. The number of articles per month broadly rose throughout the year, with peaks occurring alongside key events, such as the release of UPF research findings, books and TV shows (see **Figure 1**). Articles were published across 25 media titles in total (see **Supplementary Table 3**), with the greatest number from the Daily Mail (*n*=33, 18%). Most of the articles were published in newspapers and magazines (77%) whilst 15% were trade press publications.

#### 3.1.1 Overall position of the articles towards the UPF concept

Overall, around half of the articles accepted the UPF concept (*n*=98, 52%), 19% (*n*=35) contested the concept and 29% (*n*=55) presented a mix of statements concerning the concept (see **Figure 1** and **Supplementary Table 3)**.

### 3.2 Framing of UPFs in UK newspaper articles

Five UPF frames were identified across the articles. Frames were distinguished by their discursive positioning towards the UPF concept (accepted, contested, mixed), where responsibility was placed in the article (e.g. individual, government, industry) and whether the solutions proposed were aimed at reducing UPF consumption or identified alternative policy objectives. See **Figure 2** for an overview of the frames and **Supplementary Table 5-8** for a detailed description of each frame with additional quotes.

#### 3.2.1 Frame 1 – Social Justice

Approximately half of the articles (*n*=96, 51%) presented a framing of UPFs that aligned with Beauchamp’s^52^ conceptualisation of the ‘Social Justice’ framing of public health topics, which prioritises structural upstream solutions to protect the public from market interests^24,55^.

##### 3.2.1.1 Problem

This frame accepted the concept of UPF, often explicitly presenting it as an important dietary measure distinct from other measures, such as nutrient content (See quote (Q) 1.17 in **Supplementary Table 5**). The description of UPF products emphasised the artificial, industrially produced nature of the products, to the point that they were sometimes described as “*industrially-produced edible substances*” (Q1.36-38) rather than food. Attributes of UPF products, such as the ingredients, processes, taste, texture and packaging, were described negatively, suggesting that they are purposefully designed to lead to overconsumption (Q1.1-1.4). The cost, convenience and accessibility of UPFs were acknowledged as being beneficial to consumers but were also explained to be part of the pervasiveness of the products, which were described as ‘*addictive’*, ‘*ubiquitous’*, and *‘aggressively marketed*’ (Q1.5-1.8). In this way, UPFs were not just described at the product-level, but were positioned within dietary patterns and the wider food system^32^.

> *’Processes and ingredients used to manufacture ultra-processed foods are designed to create highly profitable, convenient, extremely palatable products that dislocate [sic] freshly prepared dishes,’ (The Telegraph, 18/02/2023)*

Within this frame diets high in UPFs were described as unhealthy. This was supported by citing or describing research linking UPFs to poor health outcomes, including weight gain, cancer, heart disease and an increased risk of mortality (Q1.13-1.15). The plurality of associations with different negative health outcomes were used to indicate a strong and “*growing body of evidence*” (Q1.13). Limitations in the research and in understanding the mechanisms were highlighted. However, the precautionary principle was used to counter these limitations (Q1.14-1.16). The high consumption of UPFs in the UK was frequently highlighted to emphasise the scale of the challenge to public health (Q 1.9). Vulnerable communities such as low-income groups and children, were identified as being at higher risk of UPF consumption and associated harms (Q 1.10-1.11). Consumers were presented as being unaware of the concept of UPFs and the associated health risks. It was proposed that consumers are being misled to think UPFs are healthy products (Q1.19-1.20). Therefore, the media attention on the topic was suggested to be warranted as individuals should be made aware about any risks. Indeed, this was proposed as necessary to trigger ‘*grassroots change’* from the public over their perception and consumption of UPFs (Q1.46).

> *“there’s now sufficient evidence to ring alarm bells. ‘Every study about UPF, as far as I know, has shown a negative impact on health,’ she says. ‘If you combine the consistency of that evidence with what we know about the health benefits of minimally processed food, it means it’s time to start thinking about how we can reduce UPFs in our shops” (The Telegraph, 23/10/23)*

##### 3.2.1.2 Cause

In the ‘Social justice’ frame, UPF manufacturers were explicitly identified as the cause of the problem. Presented as a bad actor, the UPF industry was described as deliberately engineering UPFs from cheap ingredients to be addictive with the primary purpose of increasing profit (Q1.22-1.26). Marketing strategies, including using health claims, were suggested to mislead consumers, especially parents, into thinking UPF products were healthy options (Q.1.24). There was an explicit emphasis that diet-related disease is not the individual consumer’s responsibility or due to poor willpower (Q1.48-1.51). Instead, it was suggested that the ubiquity of UPFs in the food environment effectively removes an individual’s freedom to make dietary choices (Q1.27-1.28). In this way, the cause of the problem was identified at a structural level. Delayed Government action on food policy^59^ was also proposed to contribute to creating this unhealthy food environment by allowing UPF companies to “[*get*] *away with*” (Q1.29) producing UPFs and marketing them as healthy foods (Q1.29-1.32). The UPF industry was also suggested to have influenced policymakers and researchers to create a more permissive policy environment (Q1.25).

> *’Often UPFs are cleverly marketed as healthy when they’re not, […] the food industry is not healthcare. It’s these companies’ job to make money and UPFs are the easiest way to do that because they’re designed to be moreish but not make you full.’ (The Times, 08/05/2023)*

##### 3.2.1.3 Solution

Within the ‘Social Justice’ frame, the central solution proposed was that government should introduce regulatory policy to limit harms from UPFs (Q1.39-1.44). Suggested policies included food taxes, front-of-package (FOP) labelling and advertising restrictions on UPFs.

> *He called for greater restrictions on the sale of UPFs, adding: ‘We need to make it less profitable for companies to sell us things that are going to harm us.’ (The Daily Telegraph, 28/08/23)*

Suggestions of individual dietary changes to reduce UPF intake were given, but were alongside calls for regulatory policy (Q1.47-1.50). Similarly, calls for more research to understand the mechanism between UPF and poor health were given alongside, rather than in place of, policy.

Furthermore, removing the UPF industry’s influence on research and policy was presented as a key solution to address UPF corporate power.

> *’We need to remove the influence of the industry […] Until the major charities that inform policy, the research groups doing dietary health research, and the doctors and scientists that write in the media stop taking money from the food industry, very little will change.’ (The Guardian, 03/09/2023)*

#### 3.2.2 Frame 2 – Lifestyle drift

A smaller proportion of articles contained the ‘Lifestyle drift’ frame (*n*=28, 15%). ‘Lifestyle drift’ is defined as “*the tendency for policy to start off recognizing the need for action on upstream social determinants of health inequalities only to drift downstream to focus largely on individual lifestyle factors*” ^51,55,60^.

##### 3.2.2.1 Problem

This frame also accepted the UPF concept and had a problem definition which was broadly similar to the ‘Social Justice’ frame. However, while this frame still legitimised the UPF concept, the focus of the articles was on UPF products, with the concept often being exemplified using a list of products (see Quotes 2.1-2.8 in **Supplementary Table 6**). Similarly, the consumption of UPFs was described as excessive and diets high in UPFs were linked to an increased risk of adverse health problems (Q2.9-2.12).

##### 3.2.2.2 Cause

Compared to the ‘Social Justice’ frame, where the cause of the problem was identified at the structural-level, the proposed cause in the ‘Lifestyle drift’ frame was focused on the individual-level. Thereby, there was less emphasis on the wider food system drivers of UPF consumption^32^. Instead, an individual’s knowledge of UPFs and their choice to avoid them were discussed (Q 2.19-2.22).

> *As the popular expression goes, ‘you are what you eat’ -and the same applies if your diet is unhealthy and high in preservatives, colourings and bulking agents. Little to no care towards your diet and certain foods consumed will impact your longevity and lead to a shorter life, research has shown. (The Mirror, 07/11/2022)*

UPFs were described as ‘hiding in plain sight’, which touches on the same ubiquity in the food environment as the ‘Social Justice’ frame. However, the focus was at the product-level, where UPFs were anthropomorphically described as ‘sneaking’ into people’s kitchens and diets (Q2.23-2.25), with no explicit discussion of the food system drivers that lead to this occurrence, including the companies that manufacture UPFs.

##### 3.2.2.3 Solution

Likewise, the solutions in the ‘Lifestyle drift’ frame had an individual rather than a structural focus. The articles mostly provided individual-level dietary advice to lower UPF intake (Q2.31-2.35). This was often presented as a list of foods to avoid or swap with less processed alternatives. The emphasis was placed on individuals needing to learn how to identify and avoid UPFs.

> *Whether it is a biscuit, chocolate bar or slice of cake you crave, here are five incredibly easy, delicious and more nutritious ways to get your fix, and a plethora of tips to help you avoid temptation. (The Daily Telegraph, 04/03/2023)*

#### 3.2.3 Frame 3 – Science report

A minority of articles (*n*=14, 7%) contained a ‘Science report’ framing. In this frame, the problem centred on reporting the details and results of UPF research, but it was distinct from other frames in that no explicit causes or solutions were proposed. This frame still accepted and therefore legitimised the UPF concept. However, while the reader may have concluded from the article that it would be advantageous to lower UPF consumption due to their health effects, this was not explicitly suggested.

#### 3.2.4 Frame 4 – UPF critique

The ‘UPF critique’ frame was one of two identified frames which contested the UPF concept, 20% of articles (*n=*38) contained this frame.

##### 3.2.4.1 Problem

The problem in the ‘UPF critique’ frame centred around criticisms of the UPF concept and the research associating UPF products with poor health (see **Supplementary Table 7** for more detail). The UPF definition was generally described with accuracy, using the Nova classification. The same attributes of UPF were mentioned, such as the ingredients, but were described with less negative emphasis than the ‘Social Justice’ frame (Q4.1-4.6).

> *’Emulsifiers are a wide range of compounds from synthetic compounds such as polysorbate-80 which has been linked to alterations in gut health through to lecithin which contains choline which has been shown in some circumstances to be beneficial to health.’ (Mail Online, 01/09/2023)*

Critiques of the concept included that the Nova classification is overly broad and imprecise, encompassing both ‘healthy’ and ‘unhealthy’ foods, based on their nutrient content (Q4.13-4.15). Limitations in the evidence base were also highlighted. These included the large proportion of observational studies, potential confounding factors not being considered (nutrient content, other social and behavioural factors), and the weak understanding of mechanisms driving the effect (Q4.8-4.12). Finally, consumers were presented as being aware of the UPF concept but feeling confused and worried about it (Q4.19-4.20).

##### 3.2.4.2 Cause

This frame refuted that UPFs have an independent negative health effect and instead suggested the problem was that discussion of the UPF concept caused unnecessary confusion for consumers. Unwarranted media attention was proposed as the cause of this problem. The wide media coverage of the topic was frequently highlighted, indicating that there was not enough scientific evidence to support the coverage (Q4.21-4.25).

Similar to the ‘Social Justice’ and ‘Lifestyle Drift’ frames, there was a general acceptance in this frame that diet-related disease is a concern for society. However, it was suggested that this is not an independent effect of UPFs, but rather a result of most UPFs being foods high in fat, salt and sugar (HFSS) (Q4.26-4.28), nutrients already known to be associated with adverse health effects.

> *’We need a food system which supports our health, but by getting consumers to worry or not worry about UPF is not tackling that issue’. (Mail Online, 01/09/2023)*

##### 3.2.4.3 Solutions

The solutions proposed within the ‘UPF critique’ frame were not centred on acting to reduce UPF consumption. They either proposed no action, arguing that the UPF concept should not be incorporated into policy because existing dietary recommendations and policies already address the most harmful UPFs (Q4.16-4.18, Q4.33-4.36), or they suggested deferring action until further research has been conducted (Q4.30-4.32). Most calls for research were non-specific, although some called for more randomised trials. Critically, in this frame calls for more research were suggested to be essential *before* policy action could occur. This contrasted with the ‘Social Justice’ frame, that proposed conducting further research *alongside* policy action. In this way, while there was scepticism around immediate action on UPFs, this position was presented as being open to changing, if supported by additional, sufficient evidence (Q4.38).

> *If good quality research reveals anything specifically dangerous about UPF ingredients that we can act on, we can consider doing so. But for the moment, if you follow the boring old everything in-moderation advice, there’s no reason to panic about ultraprocessed foods. (‘i’, 29/08/23)*

#### 3.2.5 Frame 5 – Market justice

The ‘Market Justice’ frame was the second frame where the UPF concept was contested, this frame was present in 24% (*n=*46) of articles. This frame aligns with Beauchamp’s^52^ conceptualisation of the ‘Market Justice’ framing of public health topics, which advocates against regulation of the market, frames industry as an responsible actor and places an emphasis on the individual causes of public health issues^24,55^.

##### 3.2.5.1 Problem

The problem definition in this frame was similar to that in the ‘UPF critique’ frame, as it argued against the UPF concept and questioned the strength of evidence linking UPFs to health harms (see **Supplementary Table 8** for more detail). It similarly suggested that media attention on the UPF concept was creating confusion among consumers and fostering distrust in industrial processing methods (Q5.28-5.30).

However, the ‘Market Justice’ frame extended this rebuttal to additionally propose that there are benefits to industrial food processing which could make food healthier and more affordable. The proposed benefits included increased food safety and security (preservatives, sterilisation etc.), allergen-free foods, lower sugar content (sweeteners), increased micronutrient content (fortification) and reduced environmental impacts. Indeed, UPFs were proposed as particularly important for low-income groups or busy families in attaining a healthy diet (Q5.26-2.27). This argument was often based on a conflation between processing and ultra-processing, as defined by the Nova classification. For example, baking, fermenting, fortification and sterilisation of foods were given as beneficial examples of processing, yet these are processes that would classify a product as processed, rather than ultra-processed (Q5.4-5.6)

In direct opposition to the ‘Social Justice’ frame where UPFs were described as containing hidden and unnatural elements, this frame often emphasised the normality of industrial ultra-processing methods and ingredients.

> *Take ultra-processed pasta sauce. […] The sugar content comes from the tomatoes and tomato puree – so no different from what you might make at home. The only unusual ingredient is lactic acid, which helps the sauce stay fresher for longer. (Mail Online, 06/05/23)*

There was comparatively little discussion in this frame of the burden of diet-related diseases compared to the ‘Social Justice’ frame. However, a need to address obesity and improve the healthfulness of the food supply was implicitly acknowledged as a problem.

Moreover, the UPF industry was presented as a good actor (Q5.31-5.35). It was described as solving challenges in the food system, complying with regulations and introducing their own regulatory measures. Interactions between industry, science and policy were also presented as beneficial (Q5.31).

> *The UK’s food and drink industry has complied with wave after wave of legislation, […] it is a dynamic market in which retailers and suppliers pursue varied solutions to the challenges and opportunities that feeding the nation affords, including innovation to reduce sugar, to reformulate plant bread and to make crisps healthier. (The Grocer, 08/06/2023)*

##### 3.2.5.2 Causes

Like the ‘UPF critique’ frame, unwarranted media attention was suggested as causing unnecessary anxiety and distrust around UPFs. At points, this extended to the suggestion that UPF manufacturers had been wrongly villainised by the media (Q5.41).

However, the UPF industry also identified its own role in creating this distrust (Q.42) via a lack of transparency about UPFs.

> *“But something else might also be at play here, […] a lack of consumer trust in the food industry. We are totally to blame, as the food industry, for that.” (WRBM Global Food, 27/04/2023)*

While there was also comparatively little discussion on the causes of diet-related disease compared to the ‘Social Justice’ frame, when this was discussed, there was greater emphasis on the role of personal responsibility (Q5.43-5.45). This was either expressed indirectly by suggesting that manufacturers abide by all regulations and are transparent about their products (Q5.31-35) or explicitly by stating that willpower is needed to lose weight (Q5.43).

##### 3.2.5.3 Solutions

The central problem in this frame was that the UPF concept is flawed, therefore it was suggested that the concept of classifying food by its processing level should not be introduced into dietary advice or policy (Q5.49-5.51).

Furthermore, as this frame argued that some UPFs can be beneficial, solutions to address societal problems related to diet and obesity centred around promoting some UPFs. On an individual level, choosing healthier UPFs was suggested as a way that consumers could achieve a healthy diet, especially for families and low-income groups (Q5.52-5.54). On a structural level, reformulation was proposed as a solution to improve nutrient content in the food supply (Q5.55), and processing as being essential to a transition towards a healthy, sustainable food system (Q5.8-5.13). To address the problem of reduced consumer trust, increased transparency around UPFs was proposed to bring consumers ‘*on-side*’ and reassure them about processing methods. This was necessary to prevent what was argued to be negative public health consequences if UPFs were avoided completely (Q5.55-5.57).

> *“there are very strong public health reasons to support reformulating to reduce the occurrence of these public health-sensitive nutrients in the food supply, so these should not be discounted due to fears about processing. (Mail Online, 27/09/23)*

Other alternative solutions proposed for improving diet and obesity were based on increasing education, implying that consumers did not understand how to attain a healthy diet (Q5.44, 5.57,5.60-5.64). Additionally, it was highlighted that addressing obesity was not solely the food industry’s responsibility to resolve, but also the responsibility of other industries (e.g. gaming due to physical inactivity) and government (Q5.63).

### 3.3 Actors and interest groups

Of the 116 identified actors in the data, 52% were academics, 20% represented NGOs, 9% were science & policy communicators, 8% were health professionals and 5% represented UPF industry (see **Table 2** and anonymised list in **Supplementary Table 9**). ‘Science & policy communicators’ refer to doctors, scientists and policy advocators who regularly discuss health topics in the media. Overall, when assessing actors’ interests, 33% of actors were categorised as ‘Interests: UPF industry related’, 20% as ‘Interests: not UPF industry related’ and 47% as ‘No interests apparent’. Interest groups were broken down by actor type, showing that the ‘UPF industry related’ group comprised mostly academics (45%) and NGO spokespeople (35%), with 11% from UPF industry spokespeople. The number of articles to which each actor contributed was examined, showing that a small number of actors were overrepresented across the articles. For example, the top 5 actors were quoted in 13-36 articles each (accounting for 30% of all quotations), whereas 63 actors were quoted in one article each (21% of all quotations). When interest group was considered, the greatest proportion of quotations were from ‘Interests: UPF industry related’ (41%) compared to ‘Interests: not UPF industry related’ (24%) and ‘No interests apparent’ (36%).

**Table 2.** - Summary of the actors quoted in articles, presented by their role and their interest group.

| Actor Group | Interest group |  |  |  |  |  |  |  |  | Total |  |
| --- | --- | --- | --- | --- | --- | --- | --- | --- | --- | --- | --- |
|  | Interests apparent |  |  |  |  |  | No interests apparent |  |  |  |  |
|  | UPF industry related |  |  | Not UPF industry related |  |  |  |  |  |  |  |
|  | N | R % | C % | N | R % | C % | N | R % | C % | N | C% |
| Academic | 17 | 28 | 45 | 8 | 13 | 35 | 35 | 58 | 64 | 60 | 52 |
| Government department | 1 | 33 | 3 | 0 | 0 | 0 | 2 | 67 | 4 | 3 | 3 |
| Health professional | 2 | 22 | 5 | 3 | 33 | 13 | 4 | 44 | 7 | 9 | 8 |
| Industry (/Spokesperson) | 4 | 67 | 11 | 2 | 33 | 8 | 0 | 0 | 0 | 6 | 5 |
| MP | 0 | 0 | 0 | 0 | 0 | 0 | 5 | 100 | 9 | 5 | 4 |
| NGO (/Spokesperson) | 13 | 57 | 35 | 5 | 22 | 21 | 5 | 22 | 9 | 23 | 20 |
| Science & Policy Communicator | 1 | 10 | 3 | 5 | 50 | 21 | 4 | 40 | 7 | 10 | 9 |
| Total | 38 | 33 | 100 | 23 | 20 | 100 | 55 | 47 | 100 | 116 | 100 |

N – Count of actors; R % - Row percent; C % - Column Percent; MP – Member of Parliament; NGO – Non-government organisation. Science & Policy Communicator – Those whose primary role involved discussing science, health and policy topics in the media.

#### 3.3.1 Relationship between actors’ interest and UPF frames

There was a relationship between the interest group of actors and the UPF frame with which they were associated (p <0.001, see **Figure 3**). First, actors with ‘No interests apparent’ and ‘Interests: not UPF industry related’ were more likely to be associated with a ‘Social Justice’ frame. Second, actors with ‘Interests: UPF industry related’ were overrepresented in the ‘UPF critique’ and ‘Market Justice’ frames (64% and 73% respectively). Nevertheless, all interest groups were represented within all five frames.

**Figure 3.**
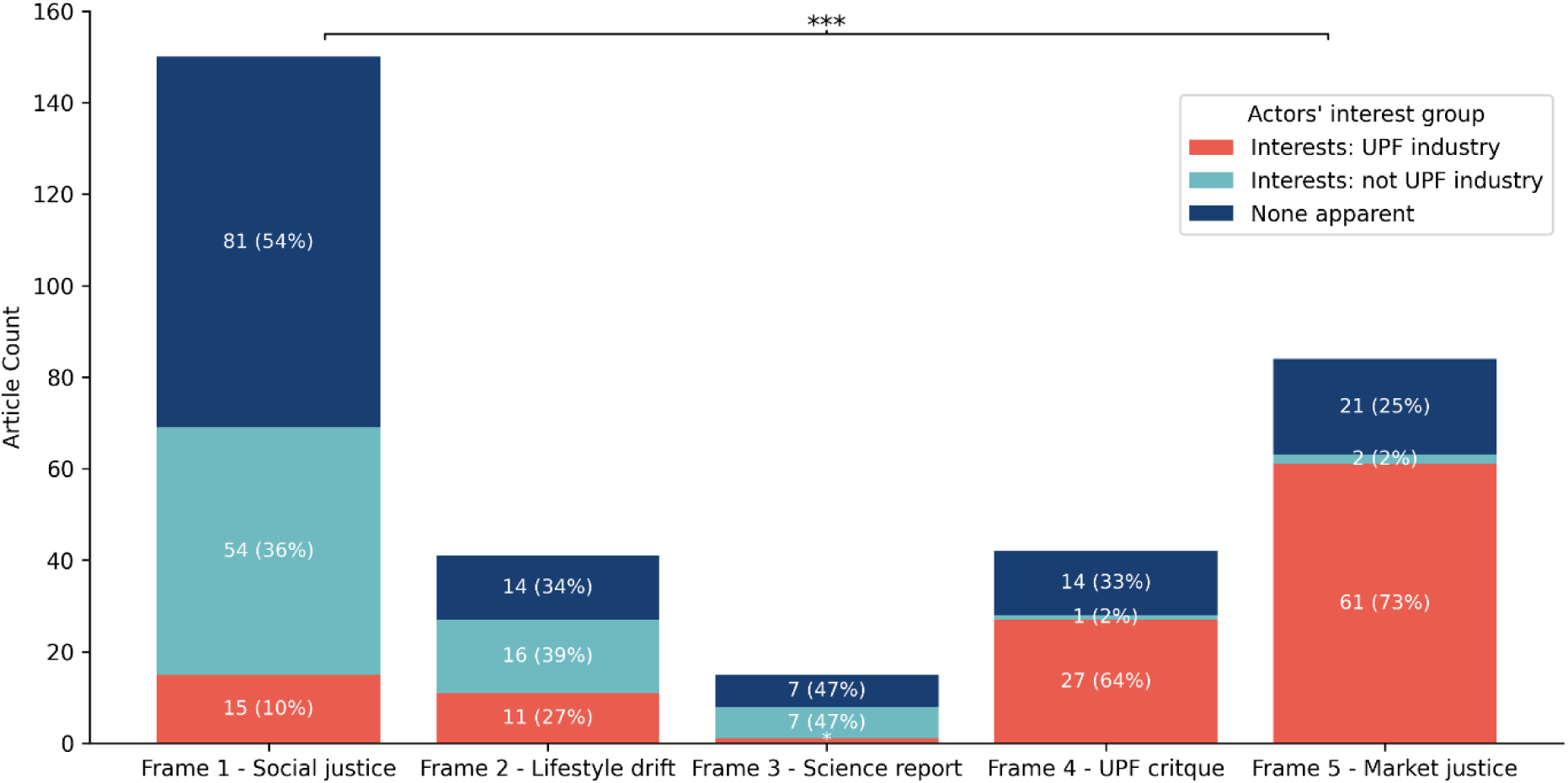
*– The frequency that actors were associated with the five UPF frames identified across UK newspaper articles (2022-23) by their interest group.* Note: *** χ² test for an actors’ interest group and association with UPF frames (p<0.001). * Frame 3, ‘Interests: UPF industry’ = 1 (7%)

## 4 Discussion

### 4.1 Key findings

In this analysis of the representation and framing of UPFs in the UK news media, we defined five frames concerning UPFs across 188 news articles published in 2022-23. A ‘Social Justice’ frame was the most prevalent (51% of articles). This highlighted the association between UPFs and poor health, emphasising the role of UPF manufacturers and the need for structural and upstream government actions to limit UPFs. In contrast, a ‘Market Justice’ frame was the second most prevalent frame (24% of articles). This questioned the concept of classifying foods by their processing level, critiqued the quality of the research associating UPFs with poor health and proposed that UPFs are essential to achieving a healthy and sustainable food system. This frame does not support regulatory policy action on UPFs but instead emphasises the importance of guiding individuals to choose healthier UPFs and industry to reformulate UPFs to improve their nutritional content.

Of the 116 actors quoted in the articles, 33% were found to have an interest related to the UPF industry. Those with UPF industry-related interests were more likely to contribute towards “Market Justice” (73%) than “Social Justice” framings of UPFs (10%). Of actors with UPF industry related interests, academics comprised the greatest proportion.

### 4.2 Relationship to prior knowledge

Only one previous study has evaluated coverage of UPFs in the news media^1^. The study examined the use of food processing terminology in the Australian media (2009 to May 2023). While there were many parallels between the studies, it is notable that our study found that 52% of articles did not contest the UPF concept compared to 85% of articles in the Australian study that did ‘*not challenge the concept […] as a measure of healthfulness*’. It is possible that the discrepancy may be due to differences between the Australian and British media and the time periods studied. However, although the definition of ‘challenge/contest’ between the studies appear to be closely aligned, it cannot be ruled out that the discrepancy was also due to methodological differences.

A greater proportion of articles that accepted the UPF concept framed it within a food systems context rather than an individual responsibility context. This represents an important break from previous media coverage of diet and obesity topics, where an individual-level framing has been persistent over time^61–64^. It will be important to investigate whether this is reflective of a broader trend in reporting of diet and obesity topics in the UK media or whether this is unique to the UPF concept, which intrinsically identifies food-system drivers. Despite this, a solely individual-level focus was still apparent in some articles, highlighting the challenge of communicating public health concepts with the public via the news media. This is analogous to the ‘Lifestyle Drift’ phenomenon^51,55,60^, whereby the wider determinants of health outcomes are initially recognised by policymakers but ultimately the proposed policies focus on individual behaviours. The message that UPF over consumption was not an individual’s responsibility in the ‘Social Justice’ frame was contradicted by articles that solely focused on individual dietary behaviours in the ‘Lifestyle Drift’ frame. In turn, this may have contributed to counterarguments that media attention on UPFs caused ‘confusion and anxiety’ in the public (as found in ‘UPF critique’ and ‘Market Justice’ frames). This is consistent with Russel *et al’s*^1^ analysis, who suggest that some of the misuse of processing terminology in the Australian media may be due to poor translation of the UPF concept from academia to news media discourse.

Many of the themes identified in the ‘UPF critique’ and ‘Market Justice’ frames in this paper resonate with well-documented corporate framing strategies deployed by harmful commodity industries, referred to as the ‘corporate playbook’^65–67^. Below we discuss how these established framing strategies resonate with themes identified in the study in greater detail.

First, Ulucanlar *et al*’s^65^ taxonomy of corporate political activity describes an overarching framing strategy whereby corporations aim to present themselves and their actions as ‘good’. Corporations place importance on maintaining a positive public image to preserve their ‘social license to operate’^68^. This is often achieved by highlighting a company’s corporate social responsibility^45,66,68^. This resonates with themes identified in the ‘Market Justice’ frame where UPF products were described as “essential” in the transition to a sustainable food system and for low-income groups. Furthermore, corporations maintain a positive image by ‘reframing discussions’ in a positive light to deflect from criticism^65,66^. In our study, UPF industry actors identified a need to reassure the public over processing methods (see Q5.57), indicating an awareness that the debate could be harming their corporate image. Garnering a positive, responsible corporate image is also a means to secure a more favourable policy environment. Harmful commodity industries often advocate for their role as ‘part of the solution’ and for voluntary over statutory measures^69^. It is therefore notable that solutions proposed under the ‘Market Justice’ frame centred around the UPF industry’s involvement, such as reformulation. Mandatory reformulation is considered to be a strong public health policy to lower specific ‘nutrients of concern’ in foods^70^. However, it has been argued that as a reformulated UPF remains a UPF, reformulation does not address the system drivers which contribute to UPF production and over-consumption^32,71^.

Second, the corporate political activity taxonomy^65^ also describes an overarching framing strategy that aims to position public health solutions and their proponents as ‘bad’. A well-documented component of this strategy is questioning the strength of scientific evidence to magnify uncertainty and manufacture doubt in public health measures^54,65,72^, which has been observed in media coverage of multiple public health topics^25,27,45^. In our study, arguments that questioned the UPF concept and strength of the scientific evidence base were present in text coded to both ‘UPF critique’ and ‘Market Justice’ frames. Importantly, the criticisms identified in these frames often represent valid scientific concerns over UPF. However, in the context of a public-facing news article where actors’ interests are obscured, it is difficult for the reader to distinguish genuine scientific debate from the deliberate use of uncertainty to undermine public health messages. However, we observed a subtle difference in frames that contested the UPF concept, which may provide insight into ways that valid scientific debate on UPFs has been utilized by the UPF industry. Whereas texts in the ‘UPF critique’ frame only critiqued the concept and science, those in the ‘Market Justice’ frame extended these critiques and argued favourably for the role of the UPF industry and its products. This could have implications for the public’s perception of the uncertainty of evidence surrounding UPFs, as Maani *et al*^54^ demonstrated that industry-sponsored messaging on health topics led to greater uncertainty on the health risks posed by industrial commodities among the public.

### 4.3 Interpretation and implications for policy and practice

This research has important implications for the communication of public health research in the news media.

First, it is notable that among actors with UPF-industry related interests in this study, only 11% were UPF industry spokespeople and the greatest proportion were academics (45%), followed by NGO spokespeople (35%). It is well established that corporations recognise the value of partnering with trusted stakeholder groups such as the scientific community^68^, including by funding scientific research. Industries are reported to use these groups to represent their positions in the media^30^, effectively distancing themselves from the message and associating it with the more trustworthy reputation of academics and NGOs. Typically, competing interests of the authors or contributors are not reported in the UK news media^73^. While academics are required to declare their interests when publishing their work in scientific journals, this information is often not present when their work reaches a public audience via the news media^73,74^. If UPF industry is going to contribute to public health debates, it is important that it is done with complete transparency to avoid misleading the public. However, it is increasingly recognised that transparency is only a partial remedy, and may have unanticipated adverse effects, and ultimately prevention of conflicts of interest is more important^75^.

Second, the presence of solely individual-focused solutions in the ‘Lifestyle Drift’ frame highlights the challenge of communicating public health concepts with the public via the news media. Researchers should work more closely with journalists to ensure balanced reporting and ensure accurate conveyance of public health messaging. This could include providing training on the difference between individual and societal drivers of disease, the nature of the commercial determinants of health, and conflicts of interest.

### 4.4 Strengths and limitations

This is the first examination of the framing of UPFs in the UK print and online news media. We used a systematic approach to identifying articles across a broad spectrum of national news media in the UK. A subsample of both the article selection and coding was performed independently by two or more researchers. Furthermore, five public members have been involved in the project throughout, providing different yet essential perspectives from the researchers.

Due to the volume of articles that mentioned UPFs, this analysis only examined articles that substantially discussed UPFs meaning it does not capture all uses of the term. Furthermore, we recognise that print and online news does not represent the total discourse on UPFs, as a declining proportion of people use print and online news sources as their main source of news, with significant generational differences^76^. However, it remains valuable as it represents the most salient discussions of the topic in the media, which is impactful on the wider debate in society^30^. The ICJME’s disclosure form was used to identify interests of those quoted in media articles. Importantly, the ICJME’s form is used in scientific and medical publishing and designed to elicit authors’ interests that might have the potential to conflict with the independence and neutrality of their scientific research. No such disclosure process routinely occurs with contributors to media content, including newspaper articles. Moreover, we recognise that some interests may have been missed in our searches as not all interests may be discernible through publicly available sources of information. Furthermore, the ICJME’s form only requires disclosures related to the previous three years^56^. Therefore, any interests older than three years will not have been captured, but may still have had an influence^77^. Whilst the most accurate date for the interest was sought, exact timings were not always available. For all these reasons, it is expected that interests were underestimated. The ICJME’s disclosure form focusses on financial interests. While we recognise that other sources of bias such as intellectual and ideological exist, it has been acknowledged that they present a ‘lower ethical priority’ than financial interests, as they are not associated with the same loss of trust^78,79^. Furthermore, as intellectual interests have a looser definition^79^, it is conceivable that nearly all actors identified in the analysis could be defined as having an intellectual interest, making it an uninformative measure in the analysis. Lastly, there are differences in the way that diet and obesity are reported in the UK media compared to other countries^80^, therefore these findings may not be transferable to other countries.

## 5 Conclusion

Public discourse concerning UPFs is rapidly growing in the UK. Globally, research is also increasing exponentially and countries across the world are now faced with decisions concerning whether to incorporate the concept of UPFs into national food policies. Media framing of UPFs could have a pivotal influence on future policy decisions and research priorities. This article sought to analyse the framing of UPFs in UK news media and trade press, finding that a ‘Social Justice’ UPF frame was dominant across the articles. This contrasts with findings from previous media analyses that reported food and obesity topics had typically been reported with an individual focus. It will be necessary to examine whether this is unique to reporting on UPFs or reflective of a wider temporal trend in journalism. However, this study also identified frames that were critical of the UPF concept. These frames were more likely to be linked to actors with UPF industry related interests, the majority of whom were academics or NGO spokespeople, rather than UPF industry spokespeople. This highlights a need for greater awareness on the interests of actors communicating public health messages to the public through the media. Whilst constructive debate is essential for high-quality science and evidence-based policy making, it is crucial that the public are aware of the provenance of all the arguments being proposed.

## Data Availability

All data are available online at Factiva

https://global.factiva.com/

## 6 Statements

## Acknowledgements

We acknowledge the involvement of five public partners throughout the work, one of whom is not a co-author on the paper (SJ).

## Author contributions

EPV, MW, JA, JCP, CM were involved in Conceptualization. JCP, EPV, JA & ME were involved in Methodology. JCP was involved in formal analysis. ME, KC, MPE, DS, MM, PS were involved in validation. EPV and JA were involved in Supervision. JCP was responsible for Writing – Original Draft Preparation. All authors were involved in Writing – Review & Editing.

## Financial Disclosure Statement

This study is funded by the National Institute for Health and Care Research (NIHR) School for Public Health Research (SPHR) (Grant Reference Number NIHR 204000). The views expressed are those of the author(s) and not necessarily those of the NIHR or the Department of Health and Social Care.

The National Institute for Health and Care Research (NIHR) School for Public Health Research (SPHR), is a partnership between the Universities of Bristol, Cambridge, Exeter and Sheffield; Imperial College London; The London School for Hygiene and Tropical Medicine; the LiLaC collaboration between the Universities of Liverpool and Lancaster; Fuse, The Centre for Translational Research in Public Health, a collaboration between Newcastle, Durham, Northumbria, Sunderland, Cumbria and Teesside Universities; and PHRESH, the Public Health RESearch for Health consortium, a collaboration between the Universities of Birmingham, Warwick and Keele.

JA, ME and MW were supported by the Medical Research Council [grant number MC_UU_00006/7]

## Supporting information

Supplementary Figure 1 – Flow diagram of article selection

Supplementary Table 1 – Description of codes for which describe the discursive position towards the UPF concept.

Supplementary Table 2 – Summary of search terms used in Google to research actor’s interests

Supplementary Table 3 – Overall article position in relation to UPFs by newspaper group and media type

Supplementary Table 4 – List of UPF corporations defined as UPF industry in the analysis

Supplementary Table 5 – Summary of the ‘Social Justice’ framing of UPF

Supplementary Table 6 – Summary of the ‘Lifestyle drift’ framing of UPF

Supplementary Table 7 – Summary of the ‘UPF critique’ framing of UPF

Supplementary Table 8 – Summary of the ‘Market Justice’ framing of UPF

Supplementary Table 9 – Anonymised list of actors present in the data and a description of their Interests

