## Supplementary Table for "Framing of ultra-processed foods and associations with interests of actors quoted in UK news media, 2022-2023: a mixed-methods study"

**Title**

**Table of figure and tables**

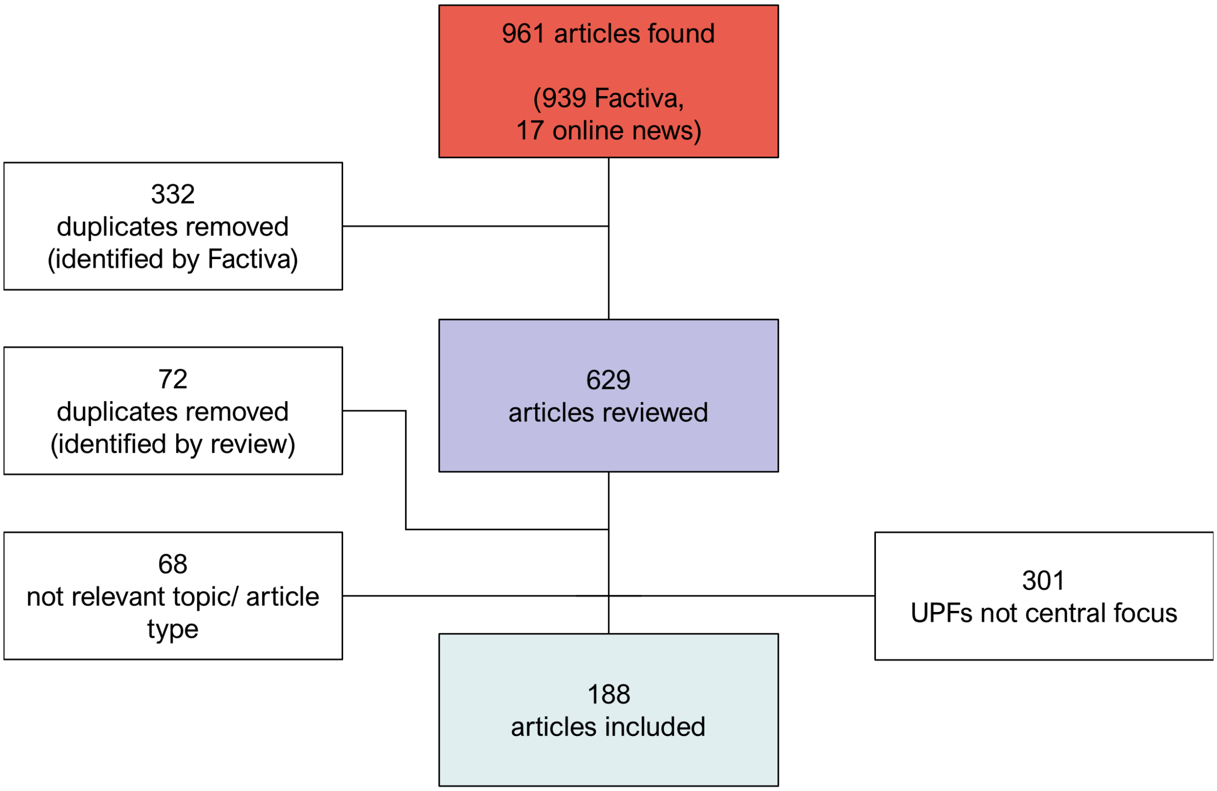

*Supplementary Figure 1 – Flow diagram of article selection*

*Supplementary Table 1 -Description of codes for which describe the discursive position towards the UPF concept.*

| Code | Description |
|---|---|
| UPF concept accepted | A statement or argument which implicitly or explicitly adopts the concept of classifying foods by their processing level.<br><br>This could include explicitly discussing the concept itself. The article may introduce defining foods by their processing level as a new, important dietary measure, discuss the merits of this approach and differences to defining food by just nutrient content.<br><br>However, it may include implicit adoption of the concept. For example, reporting on research that uses the classification to explore the associations between UPF and health, without discussing limitations of this measure or research. It could also include discussing what policy action needs to occur to reduce UPF consumption. In these instances, the concept is implicitly adopted as a measure of dietary healthfulness.<br><br>Statements where the UPF concept is adopted would lead the reader to think that the UPF concept is important, it has an independent impact on health (beyond nutrients, previous dietary advice) and/or action needs to be taken in this area, such as more research, policy action or the public changing their diet to reduce UPFs.<br><br>See example:<br><br>"*'ultra-processed foods' are classed as unhealthy, based on more than the content of salt, fat and sugar. A large body of evidence now shows ultra-processed food consumption is associated with poorer human health (including rates of heart diseases, diabetes and obesity) and planetary health (plastic pollution, excessive energy and land use, biodiversity loss).*" |
| UPF concept contested | A statement or argument which implicitly or explicitly contests the concept of classifying foods by their processing level.<br><br>These statements could include a critical discussion of or doubts in classifying foods by processing level. This may extend to discussing doubts in the quality of research on UPFs, based on the unsuitability of using processing level as a measure of dietary healthfulness.<br><br>Contesting the concept of classifying foods by their processing level may include a comparison to or preference for other measures of dietary quality, such as nutrient profile (High Fat Salt Sugar (HFSS) foods). They may also provide alternative explanations for the observed associations between UPFs and poor health outcomes, therefore questioning the urgency or importance of action in this area.<br><br>Statements which contest the UPF concept may argue for the current status quo of current dietary advice and policy. They could also indicate that people do not need to change their diets specifically to lower UPF intake. |

| | |
|---|---|
| | Statements which contest the UPF concept would lead to the reader to question the concept and therefore question the impact or importance of UPF research and may lead to the conclusion that no action needs to be taken in this area.<br><br>See example:<br>*“The Expert said that the ultra-processed foods definition is 'incredibly vague' and 'needs a lot of explanation' to be understood and implemented. He said: 'One concern with all kinds of food-related messaging that declares some foods to be 'bad' is the impact it has on people with eating disorders. But the other problem - especially now with the cost-of-living-crisis - is that people avoid perfectly fine, affordable food because they believe it to be 'unhealthy'.”* |

Supplementary Table 2 – Summary of search terms used in Google to research actor's interests

| | | |
|---|---|---|
| "Actor name" | + | "Profile"<br>"Conflict of interest"<br>"Conflicts of interest"<br>"Declaration of interests"<br>"Register of interests"<br>"Competing interests"<br>"Disclosure"<br>"Funding"<br>"Ultra-processed food" |

Supplementary Table 3 – Overall article position in relation to UPFs by newspaper group and media type

| | Overall position towards UPF concept | | | | | | | |
|---|---|---|---|---|---|---|---|---|
| | UPF concept accepted | | UPF concept contested | | Mixed | | | |
| Newspaper Group | N | R% | N | R% | N | R% | Total N | Total C% |
| **News wire** | **1** | **17** | **2** | **33** | **3** | **50** | **6** | **3** |
| Press Association | 1 | 17 | 2 | 33 | 3 | 50 | 6 | 3 |
| **Newspaper / magazine** | **80** | **55** | **23** | **16** | **42** | **29** | **145** | **77** |
| Belfast Telegraph | 1 | 100 | 0 | 0 | 0 | 0 | 1 | 1 |
| Daily Mail etc. | 16 | 48 | 4 | 12 | 13 | 39 | 33 | 18 |
| Evening Standard Online | 0 | 0 | 0 | 0 | 1 | 100 | 1 | 1 |
| Express etc. | 11 | 79 | 1 | 7 | 2 | 14 | 14 | 7 |
| Financial Times | 1 | 50 | 1 | 50 | 0 | 0 | 2 | 1 |
| i | 4 | 44 | 4 | 44 | 1 | 11 | 9 | 5 |
| Independent | 5 | 71 | 1 | 14 | 1 | 14 | 7 | 4 |
| Mirror etc. | 8 | 80 | 1 | 10 | 1 | 10 | 10 | 5 |
| New Scientist Magazine | 0 | 0 | 1 | 50 | 1 | 50 | 2 | 1 |
| New Statesman | 1 | 100 | 0 | 0 | 0 | 0 | 1 | 1 |
| The Guardian etc. | 11 | 61 | 1 | 6 | 6 | 33 | 18 | 10 |
| The Scottish Sun | 2 | 67 | 1 | 33 | 0 | 0 | 3 | 2 |
| The Spectator | 0 | 0 | 0 | 0 | 1 | 100 | 1 | 1 |
| The Sun etc. | 5 | 50 | 1 | 10 | 4 | 40 | 10 | 5 |
| The Telegraph etc. | 3 | 23 | 3 | 23 | 7 | 54 | 13 | 7 |
| The Times etc. | 12 | 60 | 4 | 20 | 4 | 20 | 20 | 11 |
| **Online news** | **5** | **63** | **1** | **13** | **2** | **25** | **8** | **4** |
| BBC News Online | 0 | 0 | 0 | 0 | 1 | 100 | 1 | 1 |
| Sky News Online | 2 | 100 | 0 | 0 | 0 | 0 | 2 | 1 |
| The Conversation | 2 | 50 | 1 | 25 | 1 | 25 | 4 | 2 |
| Yahoo News Online | 1 | 100 | 0 | 0 | 0 | 0 | 1 | 1 |
| **Trade press** | **12** | **41** | **9** | **31** | **8** | **28** | **29** | **15** |
| Dow Jones & Company, Inc. | 3 | 100 | 0 | 0 | 0 | 0 | 3 | 2 |
| Just-Food | 1 | 100 | 0 | 0 | 0 | 0 | 1 | 1 |
| The Grocer etc. | 6 | 50 | 3 | 25 | 3 | 25 | 12 | 6 |
| WRBM Global Food | 2 | 15 | 6 | 46 | 5 | 38 | 13 | 7 |
| **Grand Total** | **98** | **52** | **35** | **19** | **55** | **29** | **188** | **100** |

N – Count of articles; R % - Row percent; C % - Column Percent; ‘etc.’ – includes daily, Sunday, and online versions of the same newspaper.

Supplementary Table 4 – List of UPF corporations defined as UPF industry in the analysis

| Company Name | Definition |
|---|---|
| Arla | Top 150 global UPF corporation[1] |
| Associated British Foods | Top 150 global UPF corporation[1] |
| Cargill | Top 150 global UPF corporation[1] |
| Coca Cola | Top 150 global UPF corporation[1] |
| Danone | Top 150 global UPF corporation[1] |
| Dash Brands Ltd | Top 10 company share of Lactose Free Soft Drinks market in UK (2023) [2] |
| Ferrero | Top 150 global UPF corporation[1] |
| FrieslandCampina | Top 150 global UPF corporation[1] |
| GB Foods SA | Top 150 global UPF corporation[1] |
| General Mills | Top 150 global UPF corporation[1] |
| Hain Celestial Group Inc. (Ella's Kitchen) | Top 10 company share of Baby Food market in UK (2024) |
| Hero Group GmbH (Organix) | Top 150 global UPF corporation[1] |
| Hipp UK | Top 10 company share of Baby Food market in UK (2024) |
| Huel | Top 10 company share of High Protein Soft Drinks market in UK (2024) [2] |
| Kids Food Co Ltd (Kiddylicious) | Top 10 company share of Baby Food market in UK (2024) |
| Mars | Top 150 global UPF corporation[1] |
| Mondelez International | Top 150 global UPF corporation[1] |
| Nestlé | Top 150 global UPF corporation[1] |
| Nomad Foods Europe | Top 150 global UPF corporation[1] |
| Pagen (Pågengruppen AB) | Top 10 brand share of Baked Goods market in Western Europe (2023) [2] |
| PepsiCo | Top 150 global UPF corporation[1] |
| Pladis | Top 150 global UPF corporation[1] |
| Saputo | Top 150 global UPF corporation[1] |
| Unilever | Top 150 global UPF corporation[1] |

*Supplementary Table 5 – Summary of the 'Social Justice' framing of UPF*

<table>
<tr><th colspan="2">Frame 1 – Social Justice frame</th><th>Example quotes</th></tr>
<tr><td>Problem definition</td><td>UPF description & attributes<br><br>- UPFs contain artificial ingredients – which are unnatural, potentially harmful, disguise base ingredients and drive overconsumption.<br>- UPFs have undergone industrial processing methods which cannot be done at home, are not traditional and primarily done to increase profits.<br>- UPFs are often high in fat, salt & sugar (HFSS) they are also low in fibre and micronutrients which are lost during processing.<br>- UPFs often have a soft texture – industrial processing leads to products which are soft and easy to eat.<br>- UPFs taste appealing – they are 'hyper-palatable' leading towards overconsumption or addition.<br>- UPFs are affordable, convenient and accessible – therefore drives overconsumption, especially in low-income communities. Hard to avoid due to ubiquity.<br>- UPFs are aggressively marketed, in attractive plastic packaging. It includes food marketed as healthy using health claims. The packaging may introduce contaminants to the food<br>- UPFs are bad for the environment due to plastic pollution, reduced biodiversity, and carbon emissions<br><br>These attributes mean UPFs are unhealthy.</td><td>1.1 Ultra-processed foods (UPFs) are made using industrial processes including splitting wholefoods into oils, fats and sugar then recombining them. They are usually packed with preservatives, bought ready to eat and heavily marketed, often as healthy options. (The Times, 31/01/23)<br><br>1.2 But the lack of nutrients is not the only issue: In order to prolong their shelf life, highly processed food often contains additives such as flavor enhancers and sweeteners as well as industrial chemicals, some of which are introduced during the production process and some that leach into the food from packaging. (Barron's Online, 07/02/23)<br><br>1.3 Firstly, eating ultra-processed food makes you put on weight as it is deliberately designed to be dry and soft. He says: 'The dryness means it's high in energy and the softness means you consume it quickly. So, you consume more calories per minute than you do of real food. (The Daily Mirror, 31/08/23)<br><br>1.4 UPFs are ultra-processed foods, engineered to be as hard to resist as the TikTok algorithm: nutritionally empty, stuffed with texture and flavour enhancers. (The Guardian, 11/06/23)<br><br>1.5 Unfortunately, UPF tend to be more affordable than fresh, whole foods. They have a longer shelf life, require no preparation and can be enticing due to high sugar content that trigger feel-good dopamine responses. (The Conversation, 24/09/23)<br><br>1.6 Looking for a UPF in the typical kitchen cupboard can be like discovering an ant's nest: first you see one, then another, and before you know it, they are everywhere you look. (The Guardian, 27/08/23)<br><br>1.7 'Much of it will be familiar as 'junk food', but there's plenty of organic, free-range, 'ethical' UPF which might be sold as healthy, nutritious, environmentally friendly or useful for weight loss. Almost every food that comes with a health claim on the packet is UPF. (The Guardian, 28/08/23)<br><br>1.8 UPFs rely on energy-intensive manufacturing processes and long supply chains, leading to substantial greenhouse gas emissions. The most</td></tr>
</table>

| | | |
|---|---|---|
| | | *substantial environmental impacts of UPF-rich diets predominantly stem from the post-farm stages, specifically the final product creation and packaging processes. (The Conversation, 24/09/23*<br><br>1.9 *Yet, as he pointed out, if it's a UPF then there is no healthy version. (Mail Online, 30/04/2023)* |
| | **Consumption of UPFs**<br><br>- **Consumption of UPFs is too high** and has increased over time.<br><br>- **Children and low-income communities** as consumers at high risk, presented as victims | 1.10 *The average family's shopping basket contained less than 20 per cent ultraprocessed food in the 1970s when I was growing up. Today, it's more than 50 per cent, with poorer parts of the country at more than 80 per cent. (The Daily Mail, 22/02/23)*<br><br>1.11 *Britons lead Europe when it comes to ultra-processed foods, with the average adult consuming half of their daily calories from these foods – rising to 65 per cent in children. (Mail Online, 31/01/23)*<br><br>1.12 *The young and those on lower incomes are at particular risk, she warned, with cheap prices and aggressive marketing making them particularly attractive. (Mail Online, 31/01/23)* |
| | **Association with health**<br><br>- Consuming **UPFs is associated with a range of negative health outcomes**<br><br>- Overall, the strength of evidence supporting this association is good.<br><br>- The **body of evidence is growing;** it is consistent across different countries and outcomes<br><br>- **Studies are mostly observational**, and mechanisms are yet to be elucidated<br><br>- However, evidence is strong enough to act, **precautionary principle** is occasionally evoked | 1.13 *'We now know [UPFs are] associated with a huge number of medical problems which include weight gain and obesity, but also inflammatory problems like Crohn's disease and ulcerative colitis, metabolic diseases such as Type 2 diabetes, heart attacks, strokes, cancers, especially breast and bowel cancer, anxiety, depression, dementia and early death from all causes. [...] The potential health dangers cannot be overstated. (The Daily Mirror, 31/08/23)*<br><br>1.14 *There now appears to be incontrovertible evidence that such a diet heightens the risk not just of diabetes and cancer but cardiovascular disease and dementia. (The Guardian, 09/06/23)*<br><br>1.15 *But [she] believes there's now sufficient evidence to ring alarm bells. 'Every study about UPF, as far as I know, has shown a negative impact on health,' she says. 'If you combine the consistency of that evidence with what we know about the health benefits of minimally processed food, it means it's time to start thinking about how we can reduce UPFs in our shops and also in our shopping baskets.' (Press Association,18/05/23)*<br><br>1.16 *'While we need more research to understand the mechanisms linking UPFs to poor health, the available evidence on the harmful effects of high UPF intakes in children is too strong to ignore. (The Guardian, 08/06/23)* |

| | | |
|---|---|---|
| | **Processing level as a dietary measure**<br><br>- Defining foods by their processing level **is important and distinct from existing dietary measures** (nutrient profile/content). | 1.17 *He cites emerging evidence that the problem with UPF isn't only that it is generally high in fat, salt and sugar: there are inherent harms to eating ingredients that have undergone complex, industrial processes and are preserved, bulked out or rendered palatable by artificial additives. (New Statesman, 03/05/23)*<br><br>1.18 *He's now constantly being asked if something is UPF. Baked beans? Jar of pesto? Wine? It's a minefield. 'Sometimes I just have to say, I don't know.' Agonising over individual products misses the point, he argues. The better question is, is it real food or – as one scientist described UPF to [him] 'an industrially produced edible substance.' (The Times, 29/04/23)* |
| | **Consumers**<br><br>- Consumers are **unaware about UPFs** and their associated health risk<br><br>- Consumers, including parents, are being **misled by companies** to think that UPFs are healthy foods.<br><br>- **Consumers need to be made aware** | 1.19 *At the same time – without most of us being aware of it – we're taking huge risks with our health. (Mail Online, 22/04/23)*<br><br>1.20 *'Awareness of what is ultra-processed food is actually fairly low but they are familiar foods in your shopping trolley. (Mail Online, 22/06/23)*<br><br>1.21 *'When every penny counts, it is near criminal that families are being misled to waste money on junk food that doesn't fill you up with anything other than health risks. (WRBM Global Food, 01/02/23)* |
| **Identify cause of the problem** | **Industry as a bad actor**<br>- **UPFs are deliberately engineered by industry** to be convenient, affordable & addictive, this is done with the **purpose of generating profit**<br>- Industry uses marketing strategies to encourage and mislead consumers into buying UPFs<br>- Industry influences researchers and policymakers | 1.22 *The food industry has created the idea that all we have to worry about is fat, salt and sugar, when it is clear that food made for profit is engineered to be hard to stop eating.' (I, 04/09/23)*<br><br>1.23 *'What everyone should remember is that the food industry is not healthcare. It's these companies' job to make money and UPFs are the easiest way to do that because they're designed to be moreish but not make you full.' (The Times, 08/05/23)*<br><br>1.24 *'I strongly believe parents are being misled by companies putting health claims on ultra-processed infant food, when in fact the food is far, far anything but healthy. (The Press Association, 17/10/2023)*<br><br>1.25 *The food industry – by recruiting compliant scientists, funding studies, pushing clever marketing messages and influencing policy – has been able to cook up a self-serving narrative that shifts the blame for the harm their products cause. It is not crisps and fizzy drinks that make us fat, we are* |

| | | |
|---|---|---|
| | | *deceived into believing, but our own shortcomings in the form of sedentary lifestyles and feeble willpower. (Financial Times, 19/05/23)*<br><br>1.26*UPF's primary purpose is profit and financial growth.' (The Guardian, 12/10/23)* |
| | **Food environment**<br>- UPFs are ubiquitous, the saturated food environment makes UPFs hard or impossible to avoid | 1.27*Walking around the supermarket, temptation is everywhere: aisle after aisle of ultra-processed foods, from crisps to chocolate (The Daily Mail, 22/02/23)*<br><br>1.28*Given that those foods interact really strongly with the reward systems of the brain, if you're getting those in your face every day when you're on the high street, at the supermarket, filling up your car with petrol, it is very difficult for individuals to make healthy choices. (The Guardian, 26/10/23)* |
| | **Lack of Government Action**<br>- The Government has not acted on UPFs<br><br>- Failure to update dietary guidance, regulate food products or enact policies to restrict sales of unhealthy foods (e.g. advertisement restrictions) mentioned as specific areas of inaction | 1.29*'There's a lack of regulation on marketing and composition which means companies are getting away with marketing these products as healthier than they really are.' (The Telegraph, 18/02/23)*<br><br>1.30*This government has no appetite for action: it has shelved the ban on two-for-one offers on foods high in salt, fat and sugar in England, and delayed a ban on advertising such foods on television before 9pm. (The Guardian, 13/08/23)*<br><br>1.31*The next generation, looking back at us, will note that governments in the 2020s faced all these problems but that it took some time before political leaders understood that they all have something in common. (The Times, 19/06/23)*<br><br>1.32*He said: 'It is a shame the Government was alerted years ago but did nothing.' (Sunday Express, 03/09/23)* |
| **Make moral judgements** | **Social justice**<br>- Health (& health services) should be protected.<br>- Government should use policy to protect public health from harms of UPFs | 1.33*what is needed is a shift in government policy – a look beyond the short-term profits for big food companies and supermarkets to a longer-term view of the health of our country. (The Grocer, 24/10/23)*<br><br>1.34*Public and health agencies need to put pressure on governments to adopt new policies and implement measures that will protect public health and the environment. (The Conversation, 24/09/23)* |

| | | |
|---|---|---|
| | | 1.35 *'Britain is particularly bad for ultra-processed food. It is storing up problems for the future. If we do nothing, a tidal wave of harm will hit the NHS.' (Mail Online, 28/08/23)* |
| | **Appeal to nature**<br>- Association of natural as always being good | 1.36 *Sadly, many people are eating fake food from the moment they get up until last thing at night. (The Daily Mail, 22/02/23)*<br><br>1.37 *Without additives, she told me, the base industrial ingredients of UPF would probably not be recognisable as food by our tongues and brains. 'It would be almost like eating dirt,' she said. (Mail Online, 22/04/23)*<br><br>1.38 *a modern nutritional landscape in which 'most of our calories come from food products containing novel, synthetic molecules, never found in nature'. (Financial Times, 19/05/23)* |
| **Suggest solutions** | **Policy needed to rebalance food systems away from UPFs**<br>Government is responsible to bringing in legislation which will limit UPF companies, they are not part of the solution. Suggestions include:<br>- Calls for specific policy action to address sales of UPF, similar to tobacco:<br> o Front-of-packing labelling, taxes, marketing restrictions,<br>- Calls for changes to government dietary guidance to include UPFs<br>- Calls for public heath campaigns to improve awareness of UPFs<br>- Calls to limit industry interference in policy and research<br>- Calls for policy to make minimally processed food more accessible and affordable | 1.39 *Although, their [UPF manufacturers'] idea of the 'right thing' usually leaves much to be desired. Which takes me back to government: with UPFs, manufacturers need a big stick, not a carrot. (The Grocer, 17/06/23)*<br><br>1.40 *he does call for government intervention: first via restriction on marketing for UPF (spearing deceptive practices throughout the book). The second is more original and arguably more contentious still, calling to stop the cosy collaboration between policymakers and the food industry. (The Grocer, 04/05/23)*<br><br>1.41 *'We need taxation on UPF, we need to ban advertising, but first and foremost we need to educate people.'(Sunday Express, 03/09/23)*<br><br>1.42 *But researchers called for warning labels to be placed on processed foods, urging people to limit their intake, and said the sugar tax should be extended to cover more processed products. (The Telegraph, 31/01/23)*<br><br>1.43 *We need to put a warning about ultra processed food in our national nutrition guidance, the NHS Eatwell Guide, alongside those that mention salt, fat and sugar. This seems like a small step but it's vital (WRBM Global Food, 12/10/23)*<br><br>1.44 *We need policies that transform our failing food systems and make healthy foods available, accessible and affordable, and address the aggressive and misleading marketing of UPFs.' (The Guardian, 08/06/23)* |

| | | |
|---|---|---|
| | **Raise public awareness of risks of UPF**<br>- through public health campaigns<br>- public have a right to know if they are at risk<br>- media attention in warranted | *1.45 If there is a slight chance that it [UPFs] makes us sick we should be aware (Just-Food, 10/10/23)*<br><br>*1.46 There should be a mass campaign against ultra-processing. Campaigners such as Jamie Oliver and Henry Dimbleby have been crying out in this wilderness for years. As long as this continues, there is no way the nation's physical and mental health will improve and no way to rescue the NHS. (The Guardian, 09/06/23)*<br><br>*1.47 He says: 'The first step is grassroots change. People need to feel angry about the food they've been fed. (The Daily Mirror, 31/08/23).* |
| | **Not solely an individual's responsibility**<br>- Explicit emphasis that it is not individual's sole responsibility to reduce UPF intake | *1.48 Boycotting UPF will undoubtedly improve your personal health. But it's a lot to expect individuals to resist a multi-billion-dollar industry determined to sell us as much processed food as possible. (New Statesman, 03/05/23)*<br><br>*1.49 All the experts agree that UPF addiction cannot be solved by individuals alone. (The Guardian, 12/10/23)*<br><br>*1.50 This is a social problem that cannot be solved by telling consumers to check product labels. (The Guardian, 13/08/23)*<br><br>*1.51 Yet regulation and packaging won't change overnight so when shopping look out for: (The 'i', 14/02/23)* |
| | **Individual-level advice to lower UPFs**<br>- Advice to lower or eliminate UPFs was given<br>- This was sometimes framed in a understanding that the food environment would make this challenging | *1.52 We can all make positive changes to our diet by choosing less processed foods.*<br><br>*1.53 What do we mean when we talk about ultra-processed foods? Many people have tried to offer simple guidelines as to how to spot and avoid them. For example: Don't eat anything your grandmother wouldn't have recognised as food.*<br><br>*1.54 'cutting [UPFs] out of your diet entirely may be a challenge, […] Instead, try to minimise the amount you eat. Learn to spot them, try alternatives and stay away from as many as possible. […] While we need to take responsibility for our food, UPFs aren't our fault. (The 'i', 14/02/23)*<br><br>*1.55 On an individual level, ideally, we would be eating a more whole food diet. We would be cooking meals from scratch. While completely excising UPF from our lives may appear an uphill task, it's crucial to remember that balance is key. (Belfast Telegraph, 08/08/2023)* |

*Supplementary Table 6 – Summary of the 'Lifestyle drift' framing of UPF*

| **Frame 2 – Lifestyle drift frame** | | ***Example quotes*** |
|---|---|---|
| **Problem definition** | **UPF description & attributes**<br><br>- **UPFs contain artificial ingredients** – which are unnatural, potentially harmful, disguise base ingredients and drive overconsumption.<br>- **UPFs have undergone industrial processing** methods which cannot be done at home, are not traditional<br>- **UPFs are often high in fat, salt & sugar**<br>- **UPFs taste appealing** – they are 'hyper-palatable' leading towards overconsumption (hard to resist).<br>- **UPFs are affordable, convenient and accessible** – Hard to avoid due to ubiquity.<br>- **UPFs are aggressively marketed**, in attractive plastic packaging, often with health claims. The packaging may introduce contaminants to the food<br>- **UPFs are unhealthy** this includes foods which may be marketed as healthy foods ("hiding in plain sight") | *2.1 'But as soon as it contains emulsifiers, sweeteners, artificial preserving agents, stabilisers or colourings it becomes ultra-processed. '(The Times, 16/06/23)*<br><br>*2.2 many ultra-processed meats like wafer thin ham, contain stabilisers. These are used to enhance or help maintain their texture, structure and make it more appetising. (Mail Online, 12/08/23)*<br><br>*2.3 It's the ultra-processed food that is doing the damage, because it's been made using industrial-scale methods and ingredients that you might not recognise and wouldn't use at home to make your dinner. (The 'i', 14/02/23)*<br><br>*2.4 Get to know the traffic-light labelling on food. High salt, sugar and fat are all indicators of UPF. (Sunday Express, 27/08/23)*<br><br>*2.5 Loaded with refined grains, artificial flavours and excessive amounts of salt and sugar, these foods can flood your sensory experience, leading to overeating and addiction. (Sunday Express, 27/08/23)*<br><br>*2.6 what's happened is that UPFs have sneakily found their way into our fridges and food cupboards as the norm in their convenient packaging, low prices – particularly in a cost of living crisis – and tastiness (The 'i', 14/02/23)*<br><br>*2.7 UPFs can get everywhere – even in so-called 'healthy food'. (Mail Online, 02/09/23)*<br><br>*2.8 But it's not just crisps and chocolate bars we need to be concerned about. Many foods we would perceive as 'healthy' are also technically ultra-processed. (Mail Online, 12/08/23)* |
| | **Consumption of UPFs**<br><br>- **Consumption of UPFs is too high**<br>- **Children and low-income communities** as consumers at high risk | *2.9 Not only does the typical adult in the UK get 56.8 per cent of their calories from UPFs, according to an article published in the BMJ in 2022, the situation is worse with youngsters: around 64 per cent of calories consumed by children at lunchtime come via UPFs. (Mail Online, 24/04/23)* |

| | | |
|---|---|---|
| | - UK is among highest consumers in Europe and worldwide | 2.10 *Brits are some of the biggest consumers of ultra-processed food in Europe – making up for over 50 per cent of a daily energy intake. (The Scottish Sun, 06/06/23)* |
| | **Association with health**<br><br>- Consuming **UPFs is associated with a range of negative health outcomes**<br><br>- There are many studies which suggest UPFs are bad for a range of health outcomes | 2.11 *The growing number of UPFs in our diet has been linked to the obesity crisis. It is also thought to play a part in our risk of developing type 2 diabetes and cardiovascular disease – and even of developing certain cancers. (Mail Online, 24/04/23)*<br><br>2.12 *The spotlight is back on ultra-processed food after more studies this week suggested that too much of it increases your risk of a potentially deadly heart attack or stroke by nearly a quarter. […] We've long known they're not great for us, but with each new revelation about the possible effects of these ultra-processed foods, or UPFs, the calls for us to consume less of them become louder and louder. (Mail Online, 02/09/23)* |
| | **Processing level as a dietary measure**<br><br>- Implicitly implied as an important measure, however the concept or distinction of defining foods by their processing level is rarely discussed in detail, beyond highlighting that UPFs also include 'healthy foods' | 2.13 *'At an individual level, I think it's also important to realise that it's not the fat, the sugar, the salt that we thought it was – it's the processing. (The Express, 04/07/23)* |
| | **Consumers**<br><br>- A mixed picture is presented:<br><br>o Some articles present consumers as aware of typical advice to avoid 'junk' food but **unaware that UPFs include a broad range of foods.**<br>o Some articles present consumers as aware they should reduce UPFs but either confused how to identify them or unable to easily reduce them | 2.14 *We may think ultra processed foods like burgers and chips are obvious to spot, but it's the 'healthy' foods on supermarket shelves that we need to be wary of as well. (The 'i', 14/02/23)*<br><br>2.15 *With confusion around what constitutes UPF – and its potential effects on our health – now is the time to become informed about what we put on our plates. (Sunday Express, 27/08/23)*<br><br>2.16 *We've long known they're not great for us, but with each new revelation about the possible effects of these ultra-processed foods, or UPFs, the calls for us to consume less of them become louder and louder. (Mail Online, 02/09/23)*<br><br>2.17<br><br>2.18 *Although many of us know which foods are better for us than others, convenience and cost often influences what we actually eat. (The Express, 29/08/23)* |

| | | |
|---|---|---|
| **Identify cause of the problem** | **Lack of knowledge on UPFs**<br><br>- High consumption of UPFs is presented as being due to consumer's lack of knowledge on how to identify or avoid UPFs (by cooking etc.)<br><br>- Whilst other factors (cost, convenience, availability) may be acknowledged as important, the focus is still on individual-level factors | 2.19 *Little to no care towards your diet and certain foods consumed will impact your longevity and lead to a shorter life, research has shown. (The Mirror, 07/11/22)*<br><br>2.20 *'The problem comes when we over-consume them. It's consumers' lack of understanding about what ultra-processed is, and lack of understanding of its context and nuances within the whole diet.' (Mail Online, 02/09/23)*<br><br>2.21 *'We should be more conscious of what we're eating […] there's such a lack of nutritional knowledge and the majority of parents I work with (around 70 per cent) feed their children predominantly UPFs because of cost.' (The 'i', 30/06/23)*<br><br>2.22 *We are now too busy to do more than grab an avocado from the supermarket, or bung a ready-made meal in the microwave. We don't learn how to cook any more; we'd rather watch a TikTok influencer make a meal than do it ourselves. […] It's no wonder that UPFs have taken over our lives, drawing us into their warm, salty, sugary, fatty, instantly comforting embrace – we let them do it. (The Telegraph Online, 28/08/23)* |
| | **UPFs hiding in plain sight**<br><br>- The ubiquity of UPFs is presented as deceptive.<br><br>- While this is presented with a nefarious tone, the deceiver is not explicitly identified. | 2.23 *The problem is they can be devilishly hard to identify, with seductive packaging often marketing them as good for you. (The Times, 08/05/23)*<br><br>2.24 *Part of the problem is that these foods are so ubiquitous that they could be creeping into our diets without us realising; even some seemingly healthy foods are technically UPFs. (Mail Online, 24/04/23)*<br><br>2.25 *'what's happened is that UPFs have sneakily found their way into our fridges and food cupboards as the norm in their convenient packaging, low prices – particularly in a cost of living crisis – and tastiness' (The 'i', 14/02/23)* |
| **Make moral judgements** | **Health as paramount / 'you are what you eat'**<br><br>- Health should be protected.<br><br>- Individuals should strive to improve their health through eating a healthy diet. | 2.26 *'One thing is clear – if you want to benefit your heart health, your gut microbiome and your brain, then put aside the ultra-processed food and pick up a plant.' (The Express, 04/07/23)*<br><br>2.27 *with researchers issuing a stark warning to those wanting to live a longer and healthier life to keep these foods to a minimum (The Mirror, 07/11/22)*<br><br>2.28 *'Everything you eat contributes to how your body functions, either positively or negatively,' she says (Sunday Express, 27/08/23)* |

| | | |
|---|---|---|
| | **Appeal to nature**<br>- Association of natural as always being good | *2.29 'It's the fact that these are fake foods. They don't contain natural ingredients.' (The Express, 04/07/23)*<br><br>*2.30 'Don't have yogurt with anything added to it that isn't totally pure. (Mail Online, 04/06/23)* |
| **Suggest solutions** | **Individuals should learn to avoid UPFs**<br><br>- Individuals are presented as having a choice, can learn to avoid the 'temptation' and gain the knowledge in spotting UPFs or 'cooking from scratch'<br><br>- Articles differ on whether the advice is to reduce or fully eliminate UPFs<br><br>- No mention of broader structural solutions to aid individuals in changing their diet<br><br>- Specific dietary advice is not consistent and sometimes confuses processing and ultra-processing. | *2.31 He said: 'Most people start the day and they've got choices. They can skip breakfast, as some people do, and just have a tea or a coffee.'Or they can say, I'm not gonna have any breakfast cereal – 95 per cent of which are ultra-processed. That would be a reasonable start. (Mail Online, 04/06/23)*<br><br>*2.32 'But it's actually fairly easy to avoid ultra-processed foods if you know what they are and understand what the alternatives are. (Mail Online, 02/09/23)*<br><br>*2.33 Whether it is a biscuit, chocolate bar or slice of cake you crave, here are five incredibly easy, delicious and more nutritious ways to get your fix, and a plethora of tips to help you avoid temptation (The Daily Telegraph, 04/03/23)*<br><br>*2.34 The professor recommended looking at the back of labels when you go grocery shopping. (The Express, 04/07/23)*<br><br>*2.35 "'cutting [UPFs] out of your diet entirely may be a challenge,' the experts tell me. Instead, try to minimise the amount you eat. Learn to spot them, try alternatives and stay away from as many as possible.' […] While we need to take responsibility for our food, UPFs aren't our fault." (The 'i', 14/02/23)* |

*Supplementary Table 7 – Summary of the 'UPF critique' framing of UPF*

| **Frame 4 – UPF critique** | | ***Example quotes*** |
|---|---|---|
| **Problem definition** | **UPF description & attributes**<br><br>Nova classification consistently used to describe UPFs<br><br>- **UPFs contain artificial ingredients** –<br>- **UPFs have undergone industrial processing**<br>- **UPFs are often high in fat, salt & sugar**<br>- **UPFs taste appealing** – they are preferred by most; this *is not the same as being addictive*<br>- **UPFs have a long shelf life, are more affordable and convenient** –<br>-<br><br>However, **these attributes don't automatically make a UPF unhealthy** | 4.1 *These are foods that, broadly speaking, come in packets, are subject to industrial processing and contain ingredients you wouldn't see in a home kitchen (but more on that definition in a moment). (The Daily Mail, 01/08/23)*<br><br>4.2 *UPFs are foods where some kind of industrial production is involved, or which include ingredients like emulsifiers, sweeteners or flavour enhancers to make them tastier (The 'i', 14/10/23).*<br><br>4.3 *These are foods that are specifically designed to last a long time, be easy to store and, most importantly, taste good. ('i', 29/08/23)*<br><br>4.4 *'We've learned, over time, that it tastes nice', [...] 'That's not the same as an addiction. That's like saying we're addicted to anything we take pleasure in.'(Mail Online, 06/05/23)*<br><br>4.5 *But packaging, pricing and advertising don't alter the nutrient content of a food [...] 'you can take completely fine, healthy foods, put them together in a factory and package them – and suddenly they become ultra-processed'. (The Express, 04/09/23)*<br><br>4.6 *However, intensive processing doesn't automatically make something unhealthy. (The Express, 04/09/23)* |
| | **Consumption of UPFs**<br><br>- The level of consumption of UPFs is not consistently discussed | 4.7 *In one article, food campaigner and Leon restaurant founder Henry Dimbleby was quoted saying that 'UPF represents 55 per cent of [the UK] diet'. He is probably referring to a 2018 study that found that 56.8 per cent of foods in a typical UK diet would count as ultra-processed, according to NOVA. ('i', 29/08/23)* |
|  | **Association with health**<br><br>- Acknowledgment of association with health but **qualified with critique of the research**<br> ○ **Alternative explanations** given: association is due to nutritional content (HFSS) not processing level | 4.8 *This question is something of a nutritional hot potato. While some experts point to research linking increased UPF intake to a growing list of modern diseases, others say the research doesn't take into account that people who tend to eat most UPFs may also have other factors which contribute to ill health, such as low income. (The Daily Mail, 01/08/23)*<br><br>4.9 *This means that although adverse health associations were consistently reported, it is 'unclear' whether these associations are due to – or* |

| | | |
|---|---|---|
| | ○ **Confounding factors** (energy intake, BMI, other behavioural/socioeconomic factors) not properly accounted for<br>○ **Observational nature** of majority of research<br><br>- Highlight **limited knowledge of mechanisms** | *independent of – the 'unhealthy' nutrient contents often typical of many UPFs, such as salt, saturated fat, or free sugars. (WRBM Global Food, 12/07/23)*<br><br>*4.10 Loads of studies look at whether people who eat UPFs are unhealthier, and - unsurprisingly – they tend to be. But the studies are almost all observational (The Sunday Times, 09/09/23)*<br><br>*4.11 'This makes it extremely difficult to identify and quantify the mechanisms linking UPF diets and poor health outcomes. The subject requires further research and refinement.' (Press Association National Newswire, 11/07/23)*<br><br>*4.12 It is possible that somewhere in the long list of emulsifiers, sweeteners, flavourings and preservatives, there is something that has a peculiarly obesogenic effect. It is pure speculation, but it is not impossible. (The Telegraph Online, 06/06/23)* |
| | **Processing level as a dietary measure**<br><br>Using processing level (mostly the Nova classification) as a dietary measure is said to be flawed:<br><br>- It has **broad, poorly defined categories**<br>- The **UPF category can include foods which are considered 'healthy'**<br>- Using UPF terminology is said to place a binary on these foods; it **'vilifies' all UPFs** as being bad.<br>- **Current dietary recommendations cover most UPFs foods**, so the measure is redundant<br>- Would be **difficult to use in research, dietary advice and/or policy** | *4.13 Part of the issue is it is tricky to define what counts as an ultra-processed food. (Press Association National Newswire, 11/07/23)*<br><br>*4.14 a general problem with ultra-processed foods [is] that some options, such as sugary soft drinks, are far worse for health than other options, including wholegrain bread. (Mail Online, 28/08/23)*<br><br>*4.15 the Nova system 'lacks the precision' to work. It questions the value of using ultra-processed foods as a definition compared to assessing whether a diet is high in fat, salt and sugar or is generally less healthy. (Mail Online, 11/07/23)*<br><br>*4.16 the possibility that the observed adverse associations with ultra-processed foods are covered by existing UK dietary recommendations mean that the evidence to date needs to be treated with caution (Press Association National Newswire, 11/07/23)*<br><br>*4.17 there are too many unknowns to justify such sweeping recommendations [UPF in national dietary guidance], according to a panel of nutrition researchers (New Scientist Magazine, 16/08/23)*<br><br>*4.18 The NOVA system […] could in its simplicity and inflexibility lead to government and industry inaction on improving the nutritional content of food. (WRBM Global Food, 28/07/23)* |

| | | |
|---|---|---|
| | **Consumers**<br><br>- Presented as **aware** about UPFs but feeling **confused.** | *4.19 Even the experts find the classifications confusing. (The Express, 04/09/23)*<br><br>*4.20 He believes the term UPF breeds confusion and guilt for the consumer. 'For those who want to do research, UPF is a useful term. But, generally, it is too broad in its definition,' he says. (The Daily Mail, 01/08/23)* |
| **Identify cause of the problem** | **Unwarranted media attention**<br><br>- UPF concept (public messaging of it) causing confusion and anxiety | *4.21 The panic over UPF, recently bolstered by an episode of BBC Panorama, is not rooted in scientific evidence […] This year, mainly thanks to [the] heavily promoted book Ultra-Processed People, fear of UPF has gone mainstream. A number of journalists have written about their struggle to keep UPFs out of their lives. (The Telegraph Online, 06/06/23)*<br><br>*4.22 But the anti-UPF blowhards go far beyond telling us to have a balanced diet and eat our greens. […]. This is not a diet. It is a glorified eating disorder for the worried well. (The Telegraph Online, 06/06/23)*<br><br>*4.23 This is a media scare, not a scientific discussion – and it relies in large part on an irrational, Luddite fear of the 'unnatural'. (The 'i', 29/08/23)*<br><br>*4.24 UPFs have been hitting the headlines regularly in recent months spurred by a plethora of articles, podcasts and programmes in mainstream media, including a recent investigative documentary on the topic on UK mainstream TV . (WRBM Global Food, 27/09/23)*<br><br>*4.25 Everybody's down on UPFs, from TV chefs and medics to MPs. It's the bogeyman of the day. But UPFs aren't primarily a problem of nutrition, but a matter, first, of language and then of politics. (Financial Times, 01/09/23)* |
| | **Certainty over rising obesity, but uncertainty over UPFs as_the cause of this problem**<br><br>Acknowledgement of diet being the cause of increasing obesity and other non-communicable diseases, but disagreement with UPFs being the cause | *4.26 There's no doubt that many people struggle with eating too much, and eating too much of the wrong kind of food: the scale of the obesity epidemic is testament to that […] But that doesn't necessarily mean we can say that food, or ultra-processed food in particular, is an addictive substance. It's very unclear whether UPFs really are doing something 'special' to your body that makes them extra unhealthy (The 'i', 14/10/23)*<br><br>*4.27 There's no doubt about the threat. UPFs will kill us in droves, through pushing up rates of obesity, cancer, heart problems and type 2 diabetes. Yet the terminology used around this reality is rubbish, the descriptions dangerously weak. (Financial Times, 01/09/23* |

| | | |
|---|---|---|
| | | 4.28 *'We need a food system which supports our health, but by getting consumers to worry or not worry about UPF is not tackling that issue,' he said. (Mail Online, 01/09/23)* |
| **Make moral judgements** | **Anti appeal to nature**<br>- Rejection of association of natural as always being good | 4.29 *Bubbling under the surface is a reactionary impulse that modernity is not to be trusted, mass consumption is vulgar and a woman's place is in the home (The Telegraph Online, 06/06/23)* |
| **Suggest solutions** | **Treat current evidence with caution**<br>- More research is needed, this should happen *in place* of action<br>- **We do not yet know enough to act** | 4.30 *But experts on the Government's Scientific Advisory Committee on Nutrition (SACN) said there are 'uncertainties around the quality of evidence available'. The academics said the observed associations are 'concerning' but called for more studies to thoroughly investigate the link. (Press Association National Newswire, 10/10/23*<br><br>4.31 *But whether the concept of 'addiction' is useful for discussing food – or just confuses the issue – is still a matter for scientific debate. It seems premature to put UPFs in the same category as cigarettes and crack cocaine (The 'i', 14/10/23)*<br><br>4.32 *Another expert […], does not think we need a warning against ultra-processed foods, instead he wants to see more research.(Mail Online, 01/09/23)* |
| | **Current policies and dietary advice are sufficient**<br>- Consume everything in moderation<br>- Focus on what to eat (e.g. fruits and vegetables), rather than on what to avoid (UPFs) | 4.33 *He said: 'Our existing policies support less consumption of many of the foods that would be classified as ultra-processed because they are high in fat, salt and sugar – we know they are a problem and that's why we regulate in the way that we do. (The Independent, 21/06/23)*<br><br>4.34 *But for the moment, if you follow the boring old everything in-moderation advice, there's no reason to panic about ultraprocessed foods. (The 'i', 29/08/23)*<br><br>4.35 *From a public health perspective, 'ultra-processed food' is a major concern. But stigmatising the term, instead of focusing on the benefits of opting for healthier alternatives, is not the answer (WRBM Global Food, 09/03/23)* ,<br><br>4.36 *Rather than 'mis-demonise' foods many already know are 'not ideal', but often 'safe go-to' products, [he] wants to make it easier for the public to eat* |

| | | |
|---|---|---|
| | | *healthier foods. 'Particularly in times of money shortages and stress.' (WRBM Global Food, 09/03/23)* |
| | **UPFs can be part of a balanced diet** | 4.37 *UPFs can be a part of that balanced diet, and many of them can be perfectly nutritious. (The 'i', 29/08/23)* |
| | **Open to future action, if evidence supports** | 4.38 *'But it's absolutely vital we take a considered and very robust approach to the emerging evidence on the question of what ultra-processing is doing 'That is exactly what we're now doing. We will not hesitate to take action if the evidence suggests that is needed.' (Independent Online, 21/06/23)*<br><br>4.39 *If good quality research reveals anything specifically dangerous about UPF ingredients that we can act on, we can consider doing so. (The 'i', 29/08/23)* |

*Supplementary Table 8 – Summary of the 'Market Justice' framing of UPF*

| **Frame 5 – Market Justice** | | ***Example quotes*** |
|---|---|---|
| **Problem definition** | **UPF description & attributes**<br><br>Healthier ultra-processed foods highlighted more frequently<br><br>- **UPFs contain artificial ingredients**, which are helpful, their specific purpose is explained (e.g. increasing shelf life)<br>- **UPFs have undergone processing,** but highlight similarity between factory and home processes, some <u>*conflation of processing and ultra-processing*</u><br>- **UPFs are often high in fat, salt & sugar** however some UPFs are good sources of desirable nutrients<br>- **UPFs taste appealing** – they are preferred by most; this <u>*is not the same as being addictive*</u><br>- **UPFs can be essential for some individuals** infant formula and 'free-from' products are highlighted<br>- **UPFs have a long shelf life, are more affordable and convenient** therefore is key to food security –<br>-<br><br>However, **these attributes mean UPFs can be healthy, not all are unhealthy.** Shifting away from UPF consumption could cause negative unintended consequences | *5.1 Anything edible made with colourings, sweeteners and preservatives automatically falls into this category, according to the Nova food classification system. (Mail Online, 11/03/23)*<br><br>*5.2 Along with fish fingers, wholegrain cereals and fruit yoghurts, [they] insisted that beans on toast is a source of 'important nutrients', as well as being 'convenient and affordable'. (Mail Online, 27/04/23)*<br><br>*5.3 It means they are made in a factory with lots of ingredients listed on the back of the packet, often containing chemicals to extend shelf life. (Mail Online, 06/05/23)*<br><br>*5.4 some processing is necessary for certain foods to become edible – such as cereals and grains (Mail Online, 27/09/23)*<br><br>*5.5 Nestlé dismissed the association between ultra-processed foods and inherently unhealthy foods. 'Food processing-e.g., cooking, baking, fermenting-has been used for centuries to convert raw ingredients into safe, nutritious, enjoyable foods. Food preparation at scale is not fundamentally different from cooking at home. (WRBM Global Food, 23/11/22)*<br><br>*5.6 'Processing for food safety and stability includes controlling water (drying, frying and freezing), thermal and non-thermal pasteurisation and sterilisation, modified atmosphere packaging, and fermentation,' (WRBM Global Food, 21/04/223)*<br><br>*5.7 Take ultra-processed pasta sauce. […] The sugar content comes from the tomatoes and tomato puree – so no different from what you might make at home. The only unusual ingredient is lactic acid, which helps the sauce stay fresher for longer. (Mail Online, 06/05/23)*<br><br>*5.8 The truth is that just because a food comes in a packet it doesn't mean it is bad for you. (Mail Online, 06/05/23)*<br><br>*5.9 'We've learned, over time, that it tastes nice,'[…]'That's not the same as an addiction. That's like saying we're addicted to anything we take pleasure in. (Mail Online, 06/05/23)'* |

| | | |
|---|---|---|
| | | *5.10 But some nutritionists and food engineers argue UPFs yield numerous benefits for human and planetary health in the form of safe, nutritious, and environmentally sustainable food for the mass market. (WRBM Global Food, 03/05/23)*<br><br>*5.11 Ultra processed foods are deemed problematic because they contain preservatives – well, preservatives serve a role ,' Matte concluded. 'They reduce the risk of toxicity and allow foods to be consumed over a longer time frame – as a result, a more balanced diet over time and less food waste. There are there are some real downsides to a radical shift away from ultra processed foods that have to be evaluated. (WRBM Global Food, 01/08/23)*<br><br>*5.12 'good processing' can limit the amount of food wasted, and by consequence, its negative impact on the environment. (WRBM Global Food, 03/05/23)*<br><br>*5.13 Ultra-processed foods are often demonised for being extremely unhealthy, but in some cases they can actually be better for you than the original product (The 'i', 28/09/23)* |
| | **Consumption of UPFs**<br><br>- The level of consumption of UPFs is not consistently discussed | *NA* |
|  | **Association with health**<br><br>- Acknowledgment of association with health is mentioned but **countered with critique of the research**<br> ○ **Alternative explanations** given: association is due to nutritional content (HFSS) not processing level<br> ○ **Confounding factors** (energy intake, BMI, other behavioural/socioeconomic factors) not properly accounted for | *5.14 There is no sound scientific validation for linking all processed food to obesity. In contrast, diets high in-e.g., high in energy, sugar, or fat-may contribute to weight gain and obesity,' the [Nestlé] spokesperson told us. (WRBM Global Food, 23/11/22)*<br><br>*5.15 Similarly, arguments that some UPFs can provide nutritional benefits to consumers do not align with research linking this food category with increased risk of cancer, dementia, or mortality. So why do nutritionists and food engineers back UPFs to improve food nutrition, safety, and sustainability? They put it down to inconsistencies associated with the most common definition of UPF: categorised by the so-called NOVA food classification system. (WRBM Global Food, 03/05/23)*<br><br>*5.16 'What that says is that yes, there is a positive association, but it's actually no stronger than the association that we have seen documented for things like* |

| | | |
|---|---|---|
| | ○ **Observational nature** of majority of research<br><br>- Highlight **limited knowledge of mechanisms** | *exercise and education level and many other factors that have been associated with obesity risk. So, it doesn't really stand out in terms of magnitude of risk.' (WRBM Global Food, 01/08/23)*<br><br>*5.17 While evidence exists that ultra processed foods – classified under category four of the NOVA food classification system – are problematic for weight gain, cardiovascular disease risk and metabolic disease risk in general, there's isn't enough to support sweeping changes to policy. (WRBM Global Food, 01/08/23)*<br><br>*5.18 'SACN is absolutely concerned about the association between highly processed foods with a range of adverse health outcomes. However, we think it's unclear whether the associations are due to the nutritional characteristics of the food or whether there's any independent effect of the processing,' (New Scientist Magazine, 04/10/23)* |
| | **Processing level as a dietary measure**<br><br>Using processing level (the Nova classification) as a dietary measure is said to be flawed:<br><br>- It has **broad, poorly defined categories**<br>- The **UPF category can include foods which are considered 'healthy'**<br>- Using UPF terminology is said to place a binary on these foods; it **'vilifies' all UPFs** as being bad.<br>- **Current dietary recommendations cover most UPFs foods**, so the measure is redundant<br>- Would be **difficult to use in research, dietary advice and/or policy** | *5.19 But it's not unhealthy because it's been made in a factory. It just so happens that a lot of these products are high in fat, sugar, salt and, importantly, calories. (Mail Online, 06/05/2023)*<br><br>*5.20 He said: 'One concern with all kinds of food-related messaging that declares some foods to be 'bad' is the impact it has on people with eating disorders. (Mail Online, 11/03/23)*<br><br>*5.21 'When we look at ultra-processed foods, that definition is variable, it's complicated and indeed it's actually unworkable. (Mail Online, 27/09/23)*<br><br>*5.22 Classification systems for foods such as NOVA are ambiguous, inconsistent and often give insufficient weight to scientific evidence on nutrition and food processing methods. (WRBM Global Food, 23/11/22)*<br><br>*5.23 took issue with the NOVA system putting pre-packaged bread in the same category as carbonated soft drinks. They are both, according to NOVA, ultra-processed. But not all products within the UPF classification are the same, […] breads and cereals are UPFs if you buy them in the supermarket, but [research] shows these foods...are beneficial for diabetes.' (WRBM Global Food, 27/04/23)* |

| | | |
|---|---|---|
| | | *5.24 It warned that there is a 'lack of agreed definition' around what foods fall into the category and concerns about its 'usefulness as a tool to identify healthier products'. (Mail Online, 27/04/23)*<br><br>*5.25 I am afraid that it might result in bad governmental decisions, and it may reduce industry's attempts to improve the formulation of their food products They [food companies] can achieve a better classification within Nutri-Score, but not within NOVA. In NOVA all UPFs are equally bad. (WRBM Global Food, 28/07/23)* |
| | **Consumers**<br><br>- Presented as **aware** about UPFs but feeling **confused and anxious**<br>- Low-income families highlighted as being reliant on UPFs | *5.26 She added: 'It's great if you can cook from scratch when you have time, but I know for me, as a working parent it's often not an option. (Mail Online, 27/04/23)*<br><br>*5.27 'People rely on processed foods for a wide number of reasons, so the bottom line would be that if we remove them from our diets, this would require a huge change in the food supply, which is really unachievable for most people, and potentially resulting in further stigmatisation, guilt, etc in those who rely on processed foods, promoting further inequalities in disadvantaged groups.' (The Guardian, 28/09/2023)*<br><br>*5.28 The warnings sparked a flurry of questions from puzzled, anxious Twitter users […] It was a perfect illustration of why telling us to steer clear of processed food can risk creating more confusion. (Mail Online, 06/05/23)*<br><br>*5.29 They warned that many poorer families did not have the luxury of being able to worry about buying 'artisanal bread'. (The Telegraph, 27/09/23)*<br><br>*5.30 'Demonising all processed foods could foster feelings of guilt and stigma around food choices, adversely impacting intake of more affordable sources of nutrients. (The Times, 26/04/23)* |
| | **Industry as a good actor**<br>- Interactions with industry are positive and beneficial<br>- Industry abides by all regulation | *5.31 The benefit of working with industry is that we can access processing technologies and understand the role of processing in a real world environment. While most of my research has been publicly funded, a small amount has been collaboratively supported by food companies to tackle specific issues around food processing and functionality. (The Guardian, 03/10/23)* |

| | | |
|---|---|---|
| | | *5.32 'Food manufacturers pay us to advise them on nutritional policies to make their food healthier. We think this is important work and we're always transparent about it.' (Mail Online, 06/05/23)*<br><br>*5.33At a consumer level, Nestlé plans to continue to display locally relevant front-of-pack nutrition labelling schemes, such as Nutri-Score, on a voluntary basis or as required by authorities. Nestlé also couples its offering with consumption guidance and services to support balanced eating, the company said. (WRBM Global Food, 23/11/22)*<br><br>*5.34'Foods and drinks specifically created for infants and young children are highly regulated and are required to adhere to strict safety and nutritional standards. These processing techniques are essential in ensuring products are safe and nutritionally appropriate for the specific needs of infants and young children.' (The Guardian, 08/06/23)*<br><br>*5.35The UK's food and drink Industry has complied with wave after wave of legislation, including recent […] it: it is a dynamic market in which retailers and suppliers pursue varied solutions to the challenges and opportunities that feeding the nation affords, including innovation to reduce sugar, to reformulate plant bread and to make crisps healthier. (The Grocer, 08/06/23)* |
| **Identify cause of the problem** | **Unwarranted media attention**<br><br>- UPF concept (public messaging of it) causing confusion and anxiety<br>- It is creating a negative consumer perception of food processing, causing consumer distrust, | *5.36Ultra-processed food (UPF) has, over the past few years, become the big bad wolf of the diet world, linked to everything from cancer to bipolar disorder. (Mail Online, 06/05/23)*<br><br>*5.37Experts on nutrition. I'm sure they're only trying to help. But their findings can be so confusing. Because no sooner has one piece of breakthrough nutritional research been published, than another piece of breakthrough nutritional research comes along to contradict it. (The Daily Telegraph, 02/02/23)*<br><br>*5.38Food processing often receives a bad press. This is largely due to modern communication systems where everyone can express opinions, many of which are inaccurate and untrue. This leaves consumers concerned and often confused. (WRBM Global Food, 21/04/23)*<br><br>*5.39It was also stressed that food processing has an image problem around negative consumer perception. (WRBM Global Food, 21/04/23)* |

| | | |
|---|---|---|
| | | 5.40 *Fresh research suggests consumers struggle to distinguish between ultra-processed and other processed foods, but want to cut back on both. Is a lack of trust at play? (WRBM Global Food, 27/04/23)*<br><br>5.41 *Many of the arguments used to bash manufacturers are lazy, boring and stuck on repeat. But that's food and drink industry critics for you, all the more so now the debate about the health impacts of processed foods is being ramped up by a new champion. (The Grocer, 08/06/23)* |
| | **Lack of industry transparency as a cause of consumer distrust of UPFs** | 5.42 *Food and beverage manufacturers work behind 'closed walls' in factories, so as not to disclose IP to competitors. But the side effect of this, is that 'consumers have no idea what is going on', he explained. 'That doesn't lead to trust. […] The names of ingredients involved in food processing is another known concern amongst consumers. This, too, can feed into a lack of consumer trust, (WRBM Global Food, 27/04/23)* |
| | **Individual personal responsibility as cause of obesity**<br><br>- Industry abides by regulations and provides individuals with the information on their products<br>- individuals do not consume their products responsibly<br>- Individuals **lack education about a healthy diet** | 5.43 *Tories are supposed to believe in personal responsibility, rather than government interference. So if we want to lose weight, we should rely on old-fashioned willpower. (The Daily Telegraph, 02/02/23)*<br><br>5.44 *UPF manufacturers in the UK do not hide the nutritional values of their products.[…] Few in the industry who make UPF would argue their foods are to be consumed in abundance. No, the message from manufacturers is these products are to be eaten (and enjoyment is also often an important factor) as part of a balanced diet. The traffic light system on packaged foods makes this very easy to understand. (The Grocer, 08/06/23)*<br><br>5.45 *However, the argument and attention should move towards educating consumers, especially children in schools, about healthy, balanced diets and exercise. UPFs may be part of the problem but they are not the problem. (The Grocer, 08/06/23)* |
| **Make moral judgements** | **Market justice / pro-capitalism**<br>- Pro-capitalism stance | 5.46 *But what about all the nasty chemicals, you might ask. Don't they hijack our appetites and ruin our insides? As demonstrated above, not all processed food contains a never-ending list of alien ingredients. And not all additives deserve a bad reputation. (Mail Online, 06/05/23)*<br><br>5.47 *We should still junk the junk tax. It's the most joyless, sanctimonious, highhanded nanny-statism. Tories are supposed to believe in personal responsibility, rather than government interference. So if we want to lose weight, we should rely on old-fashioned willpower. (The Daily Telegraph, 02/02/23)* |

| | | |
|---|---|---|
| | | 5.48 *as a general principle, I don't think we should be taxing and banning things. (Press Association National Newswire, 17/10/23)* |
| **Suggest solutions** | **Should not change dietary advice**<br>- No evidence to support change in approach<br>- A change in approach would be unfeasible and possibly have unintended consequences. | 5.49 *The current way that the UK and most other countries assess the nutritional value of foods – which is generally by how much fat, salt, sugar and calories they contain – remains the best approach for achieving a healthy diet, said a panel of scientists (New Scientist Magazine, 04/10/23)*<br><br>5.50 *Advice to avoid all ultra-processed foods would be at odds with elements of current guidance and could have an impact on wider nutrient intakes.'(Press Association National Newswire, 27/09/23)*<br><br>5.51 *"urged caution for any group looking to limit the consumption of ultra processed foods, as there a very tangible potential outcomes associated with a shift away from eating many of the foods covered by the category." (WRBM Global Food, 01/08/23)* |
| | **Choosing healthier UPFs is a way to attain a healthy diet for many**<br><br>- Focus on families and low-income groups | 5.52 *'We need to make healthy eating easier and more affordable, not more difficult and expensive. Choosing healthier processed foods is one way that can help people fit healthy eating into their lives'. (Mail Online, 27/04/23)*<br><br>5.53 *'Ultra-processed foods 'definitely have a role to play' in helping people improve their health' (Mail Online, 27/09/23)*<br><br>5.54 *'we do not need to go cold turkey on them [UPFs] – but simply have to shop smarter' (The Sun, 29/08/23)* |
| | **Industry as part of the solution / good actor**<br><br>- Industrial processing can be used to **improve the nutrient profile of foods through reformulation**<br><br>- Food processing is essential for transitioning to a **sustainable food system**<br><br>- **Increased transparency and education** to **increase consumer trust** in UPF companies, processes and ingredients | 5.55 *there are very strong public health reasons to support reformulating to reduce the occurrence of these public health-sensitive nutrients in the food supply, so these should not be discounted due to fears about processing. (Mail Online, 27/09/23)*<br><br>5.56 *the discussion at the event centred on the importance of bringing consumers on-side by informing/reassuring them about existing, as well as new and emerging processing technologies. (WRBM Global Food, 21/04/23)*<br><br>5.57 *'These commitments build on the belief that transparency is key to trust,' the company said as it announced the news. 'They are also an extension of the company's long track record of helping people enjoy a variety of foods and beverages as part of a balanced diet.' (WRBM Global Food, 23/11/22)* |

| | | |
|---|---|---|
| | - Improved reporting on the **nutritional profile of** global portfolio of food products. | *5.58 food processing is essential to enable a transition to sustainable food systems (WRBM Global Food, 03/05/2023)*<br><br>*5.59 The world's largest food manufacturer [Nestlé] is working to advance the nutritional profile of its products through a strategy leveraging both reformulation and fortification (WRBM Global Food, 23/11/22)*<br><br>*5.60 Nestlé revealed it will benchmark its food and beverages against the Australian Health Star Rating system. […] We chose HSR because it is widely recognized by investors and stakeholders. It is also used by the Access to Nutrition Index to compare food portfolios of the 25 largest F&B companies globally.' (WRBM Global Food, 23/11/22)*<br><br>*5.61 A possible, successful route to consumers could be public-good, online lectures from a university or a food research institute on the pros and cons of food processing. The lecture series could be flagged extensively on national radio in small countries or on a regional basis in large countries to alert consumers. Alternatively, a TV or radio channel could host such a lecture series. (WRBM Global Food, 21/04/23)*<br><br>*5.62 'But being transparent, and [encouraging] education through labelling or health scores [or in schools] is the only way to remediate this [lack of consumer trust] (WRBM Global Food, 27/04/23)* |
| | **Improve nutrition education** | *5.63 Industries like gaming and media must also take accountability by helping to ensure consumers are active and reduce screen time in favour of exercise. Schools and the government need to do more to educate children and consumers on food and drink, because […] the UK does have an obesity problem. However, it's not up to manufacturers to bear all the responsibility. And it's not up to the government (or the pharmaceutical industry) to provide all the answers. It's up to all of us.( The Grocer Online, 08/06/23)*<br><br>*5.64 'Increasing cooking skills is undoubtedly to be encouraged, but negative messaging could imply we have to spend more money on unprocessed foods and more time in the kitchen to prepare healthier meals completely from scratch, when this is not the case.' (The Times, 26/04/23)* |

*Supplementary Table 9 – Anonymised list of actors present in the data and a description of their Interests*

| ID | # Files | Actor group | Interest group | Description of interests |
|---|---|---|---|---|
| 001 | 1 | Academic | Interests: not UPF industry | Received research funding (non-UPF industry) |
| 002 | 7 | NGO* | Interests: UPF industry | Organisation (/employer) received funding from UPF industry |
| 003 | 3 | Industry* | Interests: UPF industry | Employee (UPF industry) |
| 004 | 3 | MP | None apparent | None apparent |
| 005 | 1 | NGO* | Interests: UPF industry | Organisation (/employer) received funding from UPF industry |
| 006 | 1 | Academic | Interests: not UPF industry | Received research funding (non-UPF food industry); Royalties from food/nutrition book |
| 007 | 5 | Academic | None apparent | Author on UPF paper/academic output |
| 008 | 1 | Industry* | Interests: UPF industry | Financial interests in a food company (UPF industry) |
| 009 | 2 | MP | None apparent | None apparent |
| 010 | 1 | NGO* | None apparent | None apparent |
| 011 | 1 | Academic | None apparent | None apparent |
| 012 | 3 | Industry* | Interests: not UPF industry | Financial interests in a food company (non UPF industry) |
| 013 | 1 | Health professional | None apparent | None apparent |
| 014 | 6 | Academic | Interests: UPF industry | Received research funding (UPF industry) |
| 015 | 1 | Academic | None apparent | Author on UPF paper/academic output |
| 016 | 2 | NGO* | Interests: not UPF industry | Organisation (/employer) received funding from food/nutrition company (non-UPF industry) |
| 017 | 2 | Academic | Interests: not UPF industry | Royalties from food/nutrition book; Media appearances relating to food/nutrition; Received consulting fees (non-UPF industry); Personal interests in nutrition company (non UPF industry) |
| 018 | 2 | MP | None apparent | None apparent |
| 019 | 1 | Academic | None apparent | None apparent |
| 020 | 3 | Academic | Interests: UPF industry | Non-paid advisor to organisation that received funding from UPF industry |
| 021 | 1 | Academic | None apparent | None apparent |
| 022 | 1 | Academic | Interests: UPF industry | Received research funding from UPF industry-funded organisation |
| 023 | 4 | NGO* | Interests: not UPF industry | Organisation (/employer) campaigns on food/nutrition |
| 024 | 1 | Academic | None apparent | None apparent |
| 025 | 1 | Academic | Interests: UPF industry | Received research funding (UPF industry); Royalties from food/nutrition book |
| 026 | 9 | Academic | None apparent | Author on UPF paper/academic output |
| 027 | 3 | Academic | None apparent | Author on UPF paper/academic output |

| | | | | |
|---|---|---|---|---|
| 028 | 2 | Science & Policy Communicator | Interests: not UPF industry | Royalties from food/nutrition book; Media appearances relating to food/nutrition |
| 029 | 13 | Academic | None apparent | Author on UPF paper/academic output |
| 030 | 12 | Science & Policy Communicator | Interests: not UPF industry | Royalties from food/nutrition book; Media appearances relating to food/nutrition |
| 031 | 14 | Academic | Interests: UPF industry | Consultant / independent advisor (UPF industry); Non-paid advisor to organisation that received funding from UPF industry |
| 032 | 9 | NGO* | Interests: UPF industry | Organisation (/employer) received funding from UPF industry |
| 033 | 2 | Academic | None apparent | Author on UPF paper/academic output |
| 034 | 1 | NGO* | Interests: UPF industry | Organisation (/employer) received funding from UPF industry |
| 035 | 1 | Academic | Interests: UPF industry | Speaker honoraria (UPF industry); Received research funding (UPF industry) |
| 036 | 5 | NGO* | Interests: UPF industry | Organisation (/employer) received funding from UPF industry |
| 037 | 1 | Academic | None apparent | None apparent |
| 038 | 2 | Health professional | None apparent | Author on UPF paper/academic output |
| 039 | 1 | Academic | Interests: not UPF industry | Received speaker fees and research funding (non UPF industry) |
| 040 | 1 | Academic | None apparent | None apparent |
| 041 | 2 | Science & Policy Communicator | None apparent | None apparent |
| 042 | 5 | Academic | None apparent | Author on UPF paper/academic output |
| 043 | 1 | NGO* | Interests: UPF industry | Previous employee of organisation that received funding from UPF industry |
| 044 | 1 | Academic | Interests: not UPF industry | Author of food/nutrition book from which they receive royalties |
| 045 | 7 | Academic | None apparent | Author on UPF paper/academic output |
| 046 | 2 | Academic | None apparent | Author on UPF paper/academic output |
| 047 | 2 | Science & Policy Communicator | Interests: not UPF industry | Royalties from food/nutrition book; Media appearances relating to food/nutrition |
| 048 | 1 | Science & Policy Communicator | None apparent | Media appearances relating to food/nutrition |

| | | | | |
|---|---|---|---|---|
| 049 | 1 | Health professional | None apparent | None apparent |
| 050 | 5 | NGO* | None apparent | None apparent |
| 051 | 1 | Academic | None apparent | None apparent |
| 052 | 1 | Academic | Interests: UPF industry | Received research funding from organisation that received funding from UPF industry; Received consulting fees (non UPF industry);Personal interests in nutrition company (non UPF industry) |
| 053 | 1 | Academic | None apparent | Author on UPF paper/academic output |
| 054 | 2 | MP | None apparent | None apparent |
| 055 | 1 | Academic | None apparent | None apparent |
| 056 | 1 | Academic | None apparent | None apparent |
| 057 | 1 | NGO* | Interests: not UPF industry | Organisation (/employer) campaigns on food/nutrition |
| 058 | 1 | Academic | Interests: UPF industry | Received research funding (UPF industry) |
| 059 | 2 | Academic | Interests: not UPF industry | Received consulting fees and research funding (non UPF industry) |
| 060 | 2 | NGO* | Interests: UPF industry | Organisation (/employer) received funding from UPF industry |
| 061 | 1 | Science & Policy Communicator | None apparent | Media appearances relating to food/nutrition |
| 062 | 35 | Science & Policy Communicator | Interests: not UPF industry | Author of food/nutrition book from which they receive royalties |
| 063 | 1 | Academic | Interests: UPF industry | None declared |
| 064 | 1 | Health professional | Interests: not UPF industry | Author of food/nutrition book from which they receive royalties |
| 065 | 4 | NGO* | Interests: not UPF industry | Funded by food/nutrition company (non-UPF industry) |
| 066 | 2 | Academic | None apparent | None apparent |
| 067 | 1 | Academic | Interests: UPF industry | Member of UPF-industry linked group, Research funded by UPF industry |
| 068 | 1 | Industry* | Interests: UPF industry | Financial interests in a food company (UPF industry) |
| 069 | 1 | Academic | Interests: not UPF industry | Royalties from food/nutrition book; Received consulting fees (non food industry) |
| 070 | 7 | Academic | Interests: UPF industry | Personal interests in nutrition company (non UPF industry); Non-paid advisor to organisation that received funding from UPF industry |
| 071 | 1 | Industry* | Interests: not UPF industry | Employee of company (non UPF industry) |
| 072 | 1 | NGO* | Interests: UPF industry | Organisation (/employer) received funding from UPF industry |
| 073 | 5 | Academic | None apparent | Author on UPF paper/academic output |

| | | | | |
|---|---|---|---|---|
| 074 | 1 | NGO* | None apparent | None apparent |
| 075 | 1 | NGO* | Interests: UPF industry | Organisation (/employer) received funding from UPF industry |
| 076 | 1 | Science & Policy Communicator | Interests: not UPF industry | Royalties from food/nutrition book; Media appearances relating to food/nutrition |
| 077 | 1 | Academic | None apparent | None apparent |
| 078 | 1 | Academic | None apparent | Author on UPF paper/academic output |
| 079 | 1 | Health professional | Interests: UPF industry | Consultant (UPF industry) |
| 080 | 2 | NGO* | None apparent | None apparent |
| 081 | 1 | MP | None apparent | None apparent |
| 082 | 1 | Academic | None apparent | None apparent |
| 083 | 3 | Academic | None apparent | None apparent |
| 084 | 3 | NGO* | Interests: UPF industry | Organisation (/employer) received funding from UPF industry |
| 085 | 1 | Academic | None apparent | Author on UPF paper/academic output |
| 086 | 13 | Academic | Interests: UPF industry | Received research funding (UPF industry); Non-paid advisor to organisation that received funding from UPF industry |
| 087 | 12 | Academic | Interests: UPF industry | Received research funding (UPF industry); Personal interests in nutrition company (non UPF industry); Royalties from food/nutrition book; Media appearances relating to food/nutrition |
| 088 | 1 | Academic | None apparent | Author on UPF paper/academic output |
| 089 | 1 | Health professional | Interests: not UPF industry | Consultant / independent advisor (non UPF industry); Media appearances relating to food/nutrition |
| 090 | 1 | Academic | Interests: UPF industry | Scientific advisory board member (UPF industry) |
| 091 | 1 | Health professional | Interests: not UPF industry | Financial interests in a food company (non UPF industry) |
| 092 | 1 | Science & Policy Communicator | Interests: UPF industry | Previous employee (UPF industry) |
| 093 | 1 | Academic | None apparent | None apparent |
| 094 | 1 | Science & Policy Communicator | None apparent | None apparent |
| 095 | 2 | Academic | None apparent | Author on UPF paper/academic output |
| 096 | 2 | Academic | None apparent | Author on UPF paper/academic output |
| 097 | 1 | Academic | None apparent | Author on UPF paper/academic output |

| | | | | |
|---|---|---|---|---|
| 098 | 4 | Academic | Interests: UPF industry | Received consulting/speaker fees and research funding (UPF industry) |
| 099 | 1 | Academic | None apparent | Author on UPF paper/academic output |
| 100 | 1 | Academic | Interests: not UPF industry | Received consulting fees (non UPF industry) |
| 101 | 1 | NGO* | None apparent | None apparent |
| 102 | 2 | Academic | Interests: UPF industry | Received research funding (UPF industry) |
| 103 | 1 | Health professional | Interests: UPF industry | Received consulting fees (UPF industry); Media appearances relating to food/nutrition |
| 104 | 1 | Industry* | Interests: UPF industry | Organisation (/employer) is an UPF industry |
| 105 | 1 | Health professional | None apparent | None apparent |
| 106 | 5 | Academic | None apparent | Author on UPF paper/academic output |
| 107 | 10 | Government department | None apparent | None apparent |
| 108 | 7 | Academic | None apparent | Author on UPF paper/academic output |
| 109 | 2 | NGO* | Interests: not UPF industry | Organisation (/employer) campaigns on food/nutrition |
| 110 | 7 | Government department | Interests: UPF industry | Organisation (/employer) received funding from UPF industry |
| 111 | 1 | Academic | Interests: UPF industry | Board member to organisation that receives funding from UPF industry |
| 112 | 1 | NGO (/Spokesperson) | Interests: UPF industry | Organisation (/employer) received funding from UPF industry |
| 113 | 1 | NGO (/Spokesperson) | Interests: UPF industry | Organisation (/employer) received funding from UPF industry |
| 114 | 1 | NGO (/Spokesperson) | Interests: UPF industry | Organisation (/employer) received funding from UPF industry |
| 115 | 1 | Academic | None apparent | None apparent |
| 116 | 1 | Government department | None apparent | None apparent |

*Including organisations and spokespeople representing the organisation
